# Cancer type-specific profiling of chromatin regulator mutations identifies recurrent and co-occurring gene sets associated with improved survival following immune checkpoint therapy

**DOI:** 10.64898/2026.09.15.26363151

**Authors:** Djansel Bukovec, Marija Gjorgjievska, Sara Kocevska, Mia Nikolova, Lina Mirkovikj, Ivan Kungulovski, Zan Mitrev, Goran Kungulovski

## Abstract

Mutations in chromatin regulator (CR) genes are frequent across human cancers and have been associated with increased tumor mutational burden (TMB) and improved response to immune checkpoint inhibitor (ICI) treatment. Here, using discovery and independent validation cohorts, we resolved these associations at the level of individual cancer types and investigated whether shared CR mutation patterns emerge across colorectal cancer, melanoma, renal cell carcinoma, head and neck squamous cell carcinoma, non-small cell lung cancer, bladder cancer, esophagogastric cancer, pancreatic adenocarcinoma, prostate cancer, and breast cancer. For each cancer type, we identified a distinct set of pro-survival CR genes, whose mutations were associated with improved survival following ICI treatment. We further identified a subset of recurrent CRs that were associated with improved ICI outcomes across many cancer types, although the magnitude of the association varied between cancers. Co-occurrence of mutations in two or more CRs was associated with further improvement in survival, while several individual CR genes showed particularly robust associations with favorable ICI outcomes. Across cancer types, CR-altered groups consistently displayed higher TMB than their corresponding CR-unaltered groups. Transcriptomics-based deconvolution further revealed cancer-type-specific differences in the tumor microenvironment associated with CR mutations. Together, these findings demonstrate both cancer-specific and recurrent patterns of CR mutations associated with improved outcomes following ICI treatment and further establish a relationship between CR alterations, increased TMB, and ICI response.

## INTRODUCTION

Cancer comprises a diverse group of diseases characterized by the progressive acquisition of biological properties that enable uncontrolled cell proliferation, evasion of cell death and immune surveillance, and ultimately invasion and metastatic dissemination (1). These cancer hallmarks arise through the accumulation of molecular alterations that disrupt the regulatory systems maintaining normal cellular homeostasis and identity. Traditionally, cancer genetics has focused primarily on alterations in oncogenes and tumor-suppressor genes, including mutations, deletions, amplifications, and chromosomal rearrangements affecting pathways involved in cell proliferation, survival, DNA repair, and intracellular signaling (2).

Large-scale cancer sequencing studies have substantially expanded this view by demonstrating that genes involved in chromatin and epigenetic regulation are also frequently altered across human malignancies (3–5). Chromatin regulators (CRs), including chromatin writers, readers, erasers, and remodelers, control the organization and functional state of chromatin and thereby influence transcription, DNA accessibility, replication, and genome stability. Genetic alterations affecting these regulators can reshape epigenetic states and transcriptional programs, contributing to tumor initiation, progression, and interactions with the surrounding cellular (6–8). Thus, alongside classical oncogenes and tumor-suppressor genes, altered chromatin regulation represents an important component of the molecular architecture of cancer.

In our previous studies (9), we demonstrated across large pan-cancer cohorts that mutations in CR genes are associated with increased tumor mutational burden (TMB) and found that CR-altered tumors were associated with improved survival following immune checkpoint inhibitor (ICI) treatment, with effects comparable to tumors harboring alterations in canonical DNA-repair pathways.

Building on our previous pan-cancer observations linking CR mutations with increased TMB and favorable outcomes following ICI therapy, we investigated whether these associations persist at the level of individual cancer types and whether recurrent patterns emerge across complementary publicly available discovery and validation cohorts comprising more than 5,300 ICI-treated and 33,000 non-ICI-treated patients. We first defined cancer-specific CR mutation profiles associated with improved survival following ICI therapy and subsequently searched for recurrent CR genes shared across cancer types. We then examined whether co-occurrence of these alterations further modifies survival and TMB, identified individual CR genes with robust cancer-specific associations, and explored whether CR-altered tumors exhibit distinct tumor-microenvironment profiles. Through this stepwise approach, we sought to distinguish cancer-specific from recurrent features of CR alterations associated with outcomes following immune checkpoint therapy.

## METHOD

### Datasets used in the study

All genomic and clinical data were obtained from cBioPortal for Cancer Genomics and AACR Project GENIE (10). The analyzed resources included The Cancer Genome Atlas (TCGA), the Memorial Sloan Kettering Immune Checkpoint Inhibitor cohort (MSK-ICI), and the MSK Cancer Harmonized Oncologic Real-world Dataset (MSK-CHORD) (11–13). The discovery analysis comprised 4,439 ICI-treated patients from the combined MSK-ICI and MSK-CHORD ICI cohorts after deduplication, 5,919 TCGA non-ICI patients across 10 cancer groups, and 21,563 MSK-CHORD non-ICI patients across five cancer groups. AACR Project GENIE served as the independent validation cohort, comprising 7,402 patients across six cancer groups, including 936 ICI-treated and 6,466 non-ICI patients. Cancer-specific sample numbers are provided in Supplementary Data.

TCGA tumors were profiled predominantly by whole-exome sequencing, whereas MSK-ICI and MSK-CHORD were analyzed using the MSK-IMPACT targeted sequencing assay, covering 341–468 cancer-associated genes depending on assay version (14). GENIE samples were profiled using institution-specific targeted next-generation sequencing panels, including MSK-IMPACT, DFCI OncoPanel (15), and other hybrid-capture or amplicon-based assays, with panel composition and genomic coverage varying across participating centers.

### General statistical analysis

Statistical analyses were performed using Python, R, Microsoft Excel, and cBioPortal. Python analyses used pandas, NumPy, SciPy, statsmodels, scikit-learn, lifelines, and Matplotlib. Two-group comparisons used two-sided Mann–Whitney U tests, whereas comparisons of three or more groups used Kruskal–Wallis tests followed, where appropriate, by Dunn’s post-hoc test. For multiple parallel hypotheses, p-values were adjusted using the Benjamini–Hochberg false discovery rate (BH-FDR) within predefined cohort- and analysis-specific testing families.

### Survival analysis

Overall survival was analyzed using Kaplan–Meier curves, two-sided log-rank tests, and IPTW-weighted Cox proportional-hazards models with robust sandwich standard errors. Hazard ratios (HRs) with 95% confidence intervals (CIs) were reported, and proportional-hazards assumptions were assessed using Schoenfeld residuals where applicable. For three-group analyses, two prespecified comparisons against a common reference were performed, and log-rank p-values were BH-FDR-adjusted; no additional multiplicity correction was applied to the prespecified single comparisons.

### Selection of relevant CR genes associated with survival upon ICI

Cancer-specific pro-survival CR genes were ranked by the alive/dead ratio among mutation carriers in ICI-treated samples only. Genes requiring ≥10 mutated cases were retained, and, when possible, the ten highest-ranking genes were retained; otherwise, the maximum number meeting this criterion was selected. Non-ICI cohorts were deliberately excluded from the filtering procedure so that the survival of the ICI-derived CR gene sets could subsequently be evaluated against non-ICI cohorts without introducing selection bias. CR genes with ≥10 mutation carriers ranked by alive/dead ratio, recurring across ≥2 cancers with median ratio >1 were prioritized, and defined as recurrent genes, with EZH2 selected among three-cancer candidates based on superior average rank. Robust individual CR associations were defined as gene–cancer pairs showing a significant favorable effect in ATT-IPTW-weighted robust Cox models in ICI-treated patients (HR < 1, P < 0.05) without a corresponding significant favorable association in matched non-ICI cohorts.

### Inverse probability of treatment weighting (IPTW)

Inverse probability of treatment weighting (IPTW) targeting the average treatment effect on the treated (ATT) was applied separately within each cancer type to reduce baseline differences between the combined MSK-ICI discovery cohort and TCGA or MSK-CHORD non-ICI cohorts. The same was done for the GENIE validation cohort. Propensity scores (PS) were estimated by logistic regression using age, sex, and metastatic status. Metastatic status was harmonized as MSK-ICI sample type (metastatic vs primary), MSK-CHORD Stage IV vs Stage I–III, and TCGA Stage IV vs Stage 0/I–III; TCGA PRAD included age and sex only because metastatic status was unavailable.

ATT weights were set to 1 for ICI-treated patients and PS/(1−PS) for non-ICI patients. Propensity scores were clipped to 0.01–0.99, and non-ICI weights truncated at the 99th percentile. Balance was assessed using standardized mean differences (SMD <0.10), weight distributions, and effective sample size, calculated as (Σw)²/Σ(w²). Weighted Cox models used robust sandwich standard errors, with proportional-hazards assumptions assessed using Schoenfeld residuals.

### TMB analysis

For analyses of tumor mutational burden (TMB), Mann–Whitney U tests were used to compare mutually exclusive CR-altered and CR-unaltered tumors. Comparisons involving three or more mutually exclusive groups were assessed using Kruskal–Wallis tests followed by Dunn’s post-hoc test. For gene-level TMB analyses, BH-FDR correction was applied separately within each cohort across all planned comparisons.

### Tumor microenvironment analysis

For tumor microenvironment analyses, RNA-seq STAR-count and STAR-TPM data were obtained from the TCGA datasets deposited at the UCSC Xena GDC resource for the evaluated cancer types. Tumor microenvironment composition was estimated from linear-scale TPM expression data using two complementary deconvolution approaches: quanTIseq, using the TIL10 signature and tumor-specific mRNA scaling, and EPIC (16,17). The ESTIMATE algorithm was additionally used to calculate immune, stromal, and combined ESTIMATE scores (18). Estimated immune-cell fractions and ESTIMATE scores were compared between CR-altered and CR-unaltered tumors within each cancer type using two-sided Wilcoxon rank-sum tests. Cliff’s delta was additionally calculated as an effect-size measure for the TME comparisons.

## RESULTS

### Cancer-specific mutations in CR genes are associated with improved survival following ICI therapy

We first assessed whether mutations in a set of 26 chromatin regulator (CR) genes were associated with survival across individual cancer types. Although this set does not encompass the full spectrum of CRs recurrently mutated in cancer, it includes many well-characterized genes commonly represented in molecular profiling panels (6,7). CR-altered tumors were defined by the presence of missense, nonsense, frameshift insertion/deletion, or splice-site/splice-region mutations in at least one of the 26 CR genes (**Supplementary Figure 1**). The ICI-treated discovery cohort was generated by combining the original MSK-ICI and MSK-CHORD ICI cohorts and is referred to throughout the manuscript as MSK-ICI. It comprised colorectal cancer (CRC), melanoma (MEL), renal cell carcinoma (RCC), head and neck squamous cell carcinoma (HNSC), non-small cell lung cancer (NSCLC), esophagogastric cancer (EGC), bladder cancer (BLCA), pancreatic adenocarcinoma (PAAD), prostate adenocarcinoma (PRAD), and breast cancer (BRCA). Corresponding TCGA cohorts were used as non-ICI comparators for all 10 cancer groups, while MSK-CHORD non-ICI comparisons were additionally performed for PAAD, PRAD, NSCLC, CRC, and BRCA.

To minimize baseline differences, IPTW was applied within each cancer type. Following weighting, 37 of 44 covariate comparisons achieved adequate balance (SMD < 0.10), including all comparisons between MSK-ICI and MSK-CHORD non-ICI. Residual imbalance in the MSK-ICI versus TCGA comparisons was restricted to age and sex in PAAD, age in PRAD, age, sex, and metastatic status in NSCLC, and age in MEL (**Supplementary Figure 2**). Unless otherwise specified, IPTW-weighted cohorts were used for subsequent analyses.

Forest-plot analysis of the 26-CR set across individual cancer types showed HRs below 1 for most ICI-treated cancers, with a median HR of 0.68 compared with 0.99 in the corresponding non-ICI cohorts, indicating better overall survival **(Figure 1A)**. We next identified cancer-specific pro-survival CR sets by ranking genes according to the alive/deceased ratio among mutation carriers in ICI-treated samples only; non-ICI cohorts were not used for selection, allowing unbiased evaluation of the ICI-derived gene sets in these comparison cohorts. Genes with ≥10 mutated cases were eligible, and up to the ten highest-ranking genes were retained. Ten genes were selected for CRC, MEL, NSCLC, and BLCA, with smaller sets for EGC (2 genes), HNSC (3 genes), RCC (4 genes), PAAD (1 gene), BRCA (7 genes), and PRAD (7 genes), resulting in lower statistical power in cancers with fewer eligible genes (Supplementary Data). These cancer-specific sets showed similar separation between ICI and non-ICI cohorts with median HRs of 0.72 and 1.01, respectively (**Figure 1B**), indicating that the favorable survival associations were largely concentrated in ICI-treated cohorts. In both analyses, individual associations were not always statistically significant, likely reflecting limited sample size and cancer-specific heterogeneity; however, the overall direction of the effects was consistent across most ICI-treated cancer types.

**Figure 1.**
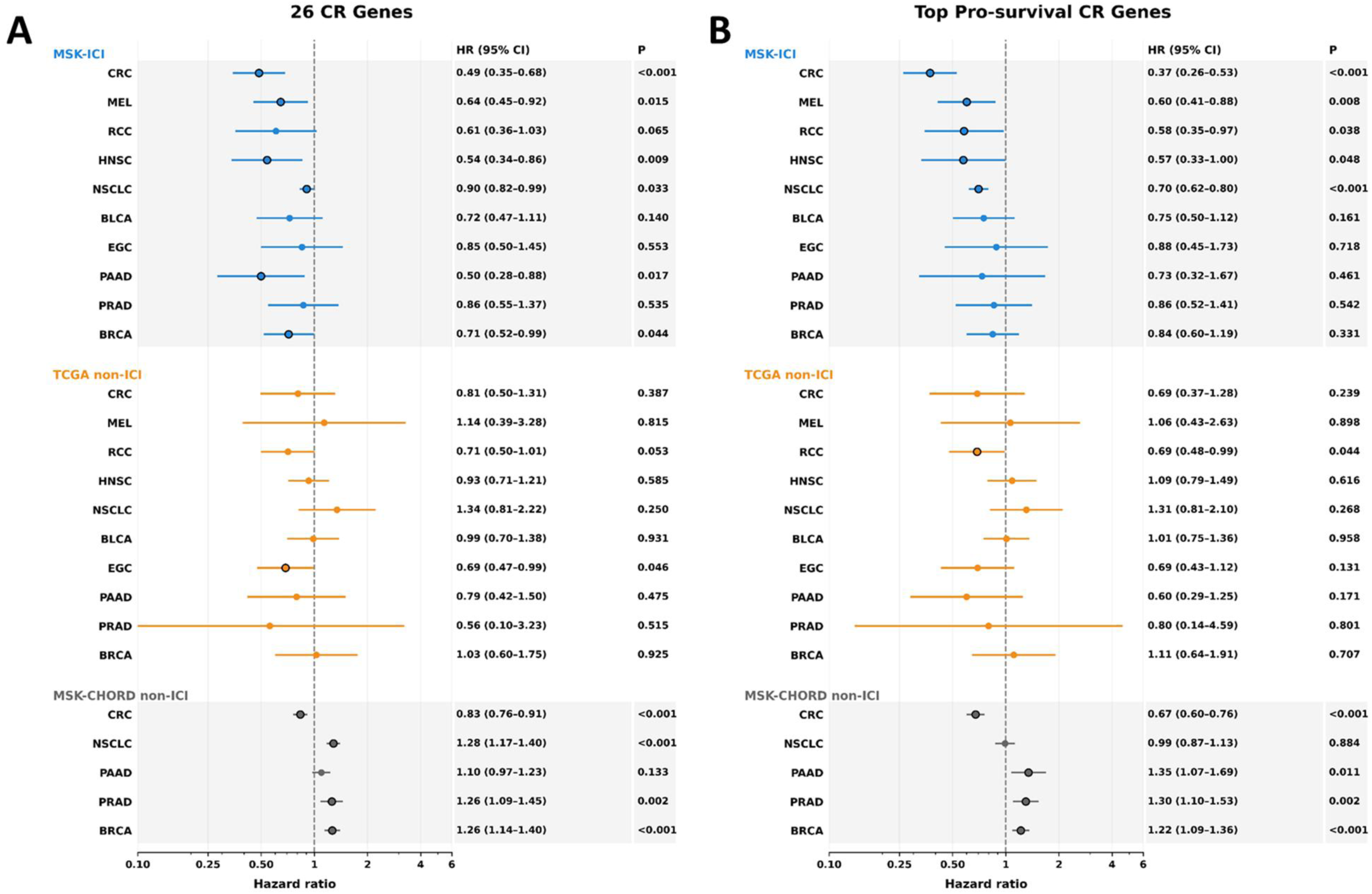
Association of chromatin regulator mutations with overall survival in ICI-treated and non-ICI-treated cohorts. Forest plots show hazard ratios (HRs) and 95% confidence intervals (CIs) associated with CR mutations across individual cancer types, allowing comparison of their survival effects between ICI-treated and non-ICI-treated cohorts. (A) Survival associations for tumors harboring mutations in any of the 26 analyzed CR genes. (B) Survival associations after restricting the analysis to cancer-specific pro-survival CR gene sets. ICI-treated cohorts comprised the original MSK-ICI cohort and the ICI-treated subset of MSK-CHORD, which were combined after deduplication and are collectively referred to as MSK-ICI throughout the manuscript; non-ICI comparisons comprise the corresponding IPTW-matched TCGA cohorts and the non-ICI subset of MSK-CHORD. HR < 1 indicates a lower mortality hazard associated with CR mutations, whereas HR > 1 indicates a higher mortality hazard. Points represent HR estimates and horizontal lines indicate 95% CIs. Exact HRs, 95% CIs, and P values are shown on the right. Bold values indicate statistically significant associations (P < 0.05). Colors denote the respective cohorts as indicated in the figure. This figure is related to Supplementary Figures 1-4. The n values for all analyzed groups can be found in Supplementary Data.

The corresponding Kaplan–Meier analyses further supported these cancer-specific survival patterns (**Supplementary Figures 3 and 4**). In the combined MSK-ICI cohort, 26 CRs-altered and pro-survival CR-altered tumors showed consistently better survival than CR-unaltered tumors across most of the evaluated cancer types, with significant associations in CRC, MEL, RCC, HNSC, and NSCLC. Other cancers showed the same general direction but did not reach statistical significance, likely reflecting smaller CR-altered subgroups and cancer-specific heterogeneity. In contrast, the corresponding TCGA non-ICI cohorts generally showed less favorable survival (**Supplementary Figure 3A and 4A**). Finally, direct comparison of MSK-ICI-treated and MSK-CHORD-non-ICI patients again showed better survival for the selected CR-altered groups in most cancer types, with significant differences in CRC and NSCLC (**Supplementary Figures 3B and 4B**). Weaker or absent associations generally occurred in cancers with smaller ICI-treated subsets, residual IPTW imbalance, and/or reduced pro-survival CR sets. Overall, these findings indicate that the survival advantage associated with the selected pro-survival CR genes was substantially more consistent in ICI-treated than in non-ICI-treated patients.

### Recurrent and co-occurring CRs associated with improved ICI outcomes across most tested cancer types with varied magnitude

We next investigated whether CR genes associated with improved survival recurred across cancer types and whether a common set of recurrent CRs could be identified. Six recurrent genes: *ARID1A, KMT2D, ARID1B, CREBBP, KMT2A, and EZH2* were selected using a composite score based on their alive/dead ratio among mutation carriers and the number of cancer types in which this ratio was favorable **(Supplementary Figure 5)**. Forest-plot analysis of this six-gene set showed a consistent shift toward lower hazard ratios in the ICI-treated cohorts, with a median HR of 0.77, compared with 1.0 in the non-ICI cohorts **(Figure 2A)**.

**Figure 2.**
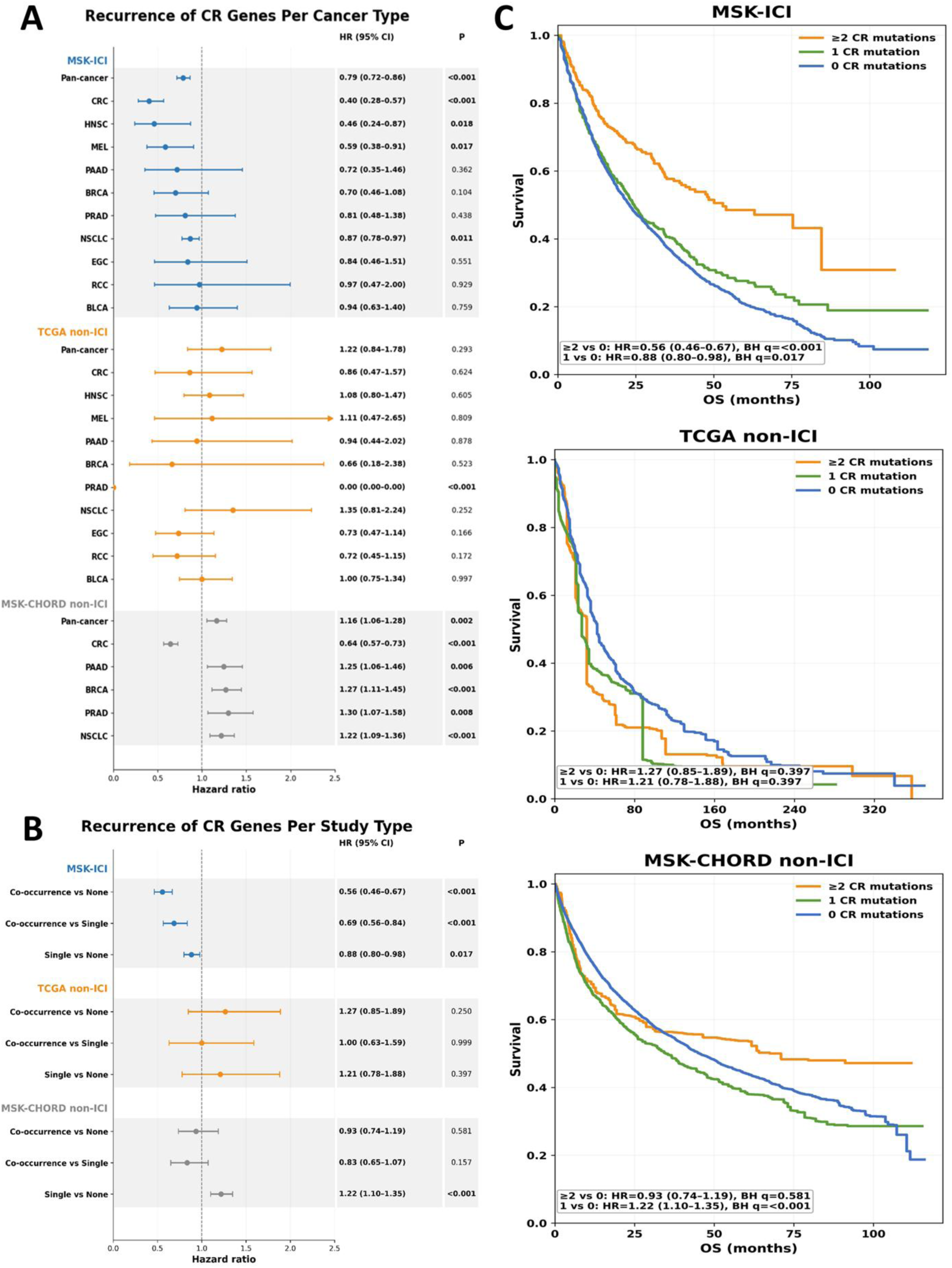
Survival associations of recurrent and co-occurring chromatin regulator mutations in ICI-treated and non-ICI-treated cohorts. (A) Forest plot showing hazard ratios (HRs) and 95% confidence intervals (CIs) associated with mutations in the six recurrent CR genes (*ARID1A, SETD2, KMT2A, KMT2D, CREBBP,* and *EZH2*) across individual cancer types and pan-cancer groups, allowing comparison of their survival effects between ICI-treated and non-ICI-treated cohorts. (B) Forest plot evaluating the effect of CR mutation co-occurrence within the six-gene recurrent set. Tumors harboring mutations in two or more recurrent CR genes (co-occurring) were compared with tumors harboring a single recurrent CR mutation or no recurrent CR mutations. (C) IPTW-weighted pan-cancer Kaplan–Meier curves showing overall survival according to recurrent CR mutation burden (≥2, 1, or 0 mutations) in MSK-ICI and matched TCGA non-ICI, and MSK-CHORD non-ICI cohorts. HR < 1 indicates lower mortality hazard for the indicated CR-altered group relative to the reference group, whereas HR > 1 indicates higher mortality hazard. Points represent HR estimates and horizontal lines indicate 95% CIs. Exact HRs, 95% CIs, and P values are shown in the forest plots, and pairwise survival statistics are shown within the Kaplan–Meier panels. Bold values indicate statistically significant associations (P < 0.05). Colors denote the respective cohorts or CR-mutation groups as indicated in the figure. This figure is related to Supplementary Figures 5-9. The n values for all analyzed groups can be found in Supplementary Data.

Cancer-specific Kaplan–Meier analyses were consistent with the forest-plot findings (**Supplementary Figure 6**). In the combined MSK-ICI cohort, recurrent CR alterations were associated with favorable survival trends across all evaluated cancer types, reaching statistical significance in CRC, HNSC, MEL, and NSCLC. Corresponding TCGA non-ICI comparisons were predominantly null, with PRAD representing the only significant favorable association (**Supplementary Figure 6A**). In comparisons with MSK-CHORD non-ICI, CRC and NSCLC retained a favorable association, whereas PAAD, BRCA, PRAD, and NSCLC showed significantly unfavorable associations in the non-ICI setting (**Supplementary Figure 6B**). Overall, recurrent CR alterations were preferentially associated with favorable survival in ICI-treated patients.

We next examined whether co-occurring CR mutations were associated with greater survival benefit. Across the complete 26-gene set, tumors with mutations in at least two CR genes showed better survival than tumors with one or no CR mutations in ICI-treated cohorts (**Supplementary Figure 7A**). Restricting the analysis to the six recurrent CRs showed the same pattern: co-occurrence of ≥2 mutations was associated with further survival improvement in ICI-treated groups, whereas non-ICI comparisons were predominantly null or unfavorable (**Figure 2B and C**). This was evident in the cancer-specific Kaplan–Meier analyses, which similarly showed better survival for ≥2 versus single recurrent CR mutations in most ICI-treated cancers, with no comparable benefit in most non-ICI cohorts; CRC in MSK-CHORD non-ICI was the main exception (**Supplementary Figure 7B and C**). These findings indicate that co-occurring CR alterations are associated with more favorable survival in ICI-treated patients.

Finally, individual CR genes with robust cancer-specific survival associations in ICI-treated but not corresponding non-ICI cohorts were identified. These included *CREBBP* in BLCA, *ARID1A* in HNSC, and *KMT2D* in MEL, and *CREBBP* in CRC and *CREBBP and KMT2A* in NSCLC (**Supplementary Figures 5, 8 and 9**).

Overall, recurrent CR mutations were associated with favorable survival across multiple ICI-treated cancers, with stronger associations when multiple recurrent CR alterations co-occurred. The weaker, absent, or occasionally unfavorable effects in non-ICI cohorts further support that these associations are preferentially observed in the context of immune checkpoint therapy.

### CR-altered tumors exhibit higher tumor mutational burden than corresponding CR-unaltered tumors in all tested cancer types

We next examined the relationship between mutations in chromatin regulator (CR) genes and tumor mutational burden (TMB). In our previous pan-cancer analysis of tens of thousands of tumor samples, CR mutations were consistently associated with elevated TMB (9). Because baseline mutational burden differs substantially among cancer types (19), we resolved this analysis at the level of individual cancers and evaluated ICI-treated and non-ICI-treated cohorts separately **(Supplementary Figure 10)**.

We first compared tumors with mutations in any of the 26 CR genes with CR-unaltered tumors. Across all evaluated cancer types and treatment cohorts, CR-altered tumors showed significantly higher TMB (**Figure 3A and B; Supplementary Figure 11,** two-sided Mann–Whitney U p < 0.001), indicating that this association persisted within individual cancers. The same pattern was observed for the cancer-specific pro-survival CR sets in both ICI-treated and non-ICI cohorts (**Figure 3C and D; Supplementary Figure 12**). Finally, analysis of the six recurrent CR genes showed a stepwise increase in TMB with mutation burden: tumors with ≥2 recurrent CR mutations had the highest TMB, followed by single-mutant and CR-unaltered tumors (**Kruskal–Wallis, p < 0.001; Supplementary Figure 13**). Overall, CR mutations were consistently associated with higher TMB across cancer types.

**Figure 3.**
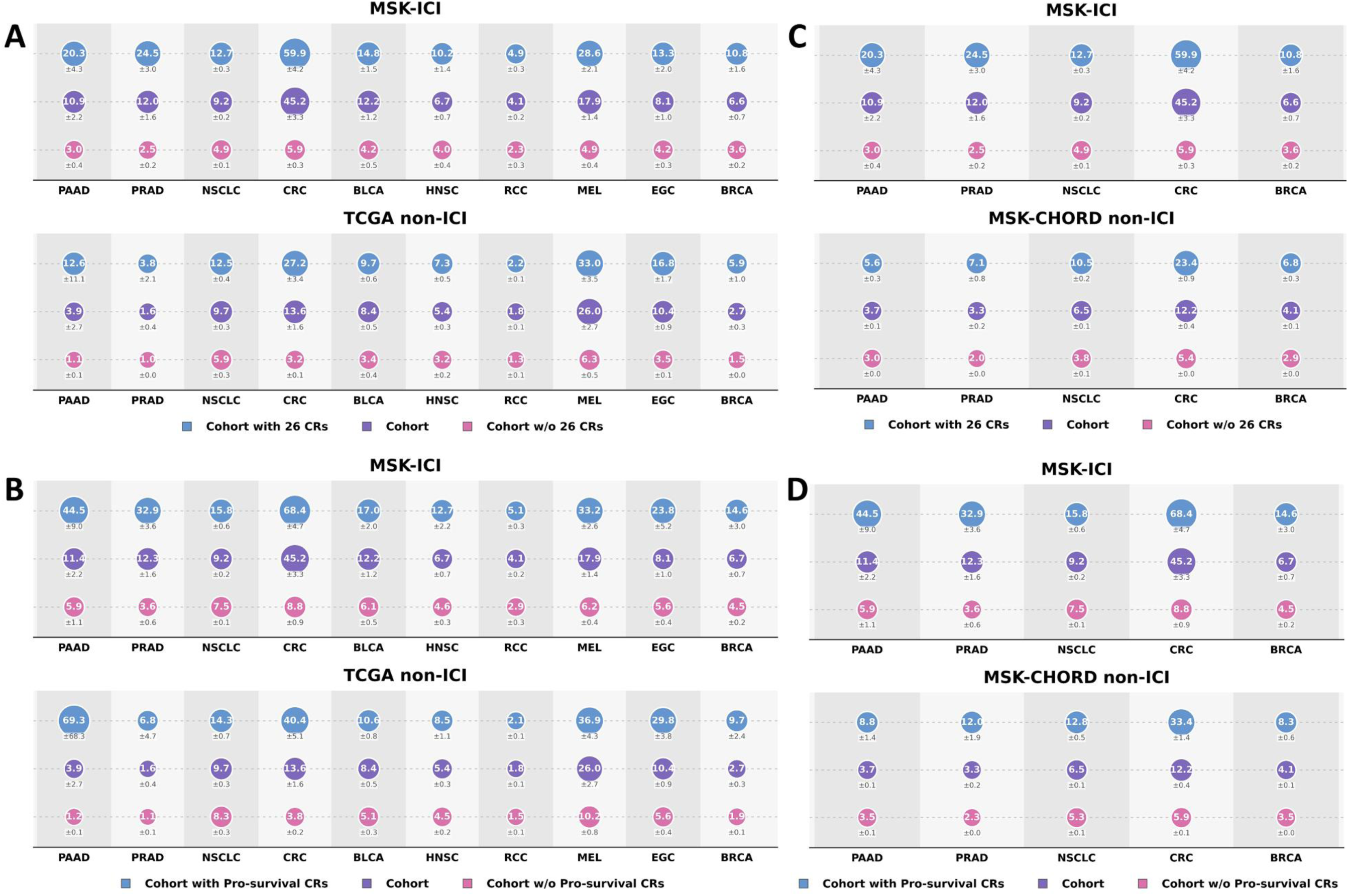
Tumor mutational burden in CR-altered tumors across ICI-treated and non-ICI-treated cohorts. Tumor mutational burden (TMB) is shown for tumors harboring mutations in the complete 26-CR gene set or the cancer-specific pro-survival CR sets, the corresponding overall cancer cohort, and tumors lacking mutations in the respective CR gene set. (A) Complete 26-CR set in the combined MSK-ICI cohort and corresponding TCGA non-ICI cohorts across 10 cancer types. (B) Complete 26-CR set in MSK-ICI and MSK-CHORD non-ICI cohorts across the five cancer types represented in both datasets. (C) Cancer-specific pro-survival CR sets in MSK-ICI and corresponding TCGA non-ICI cohorts. (D) Cancer-specific pro-survival CR sets in MSK-ICI and MSK-CHORD non-ICI cohorts. Circle size is proportional to TMB, with the corresponding TMB value shown within each circle and variability indicated below. Colors denote the respective CR-defined groups as indicated. This figure is related to Supplementary Figures 10–14. The n values for all analyzed groups can be found in Supplementary Data.

To assess the contribution of TMB to the observed survival associations, we compared unadjusted and TMB-adjusted Cox proportional-hazards models for both cancer-specific pro-survival CR sets and co-occurring recurrent CR mutations. The impact of TMB adjustment varied across cancer types, with most of the associations becoming attenuated or no longer statistically significant, but some of them remaining largely preserved even after the TMB adjustment. This heterogeneity indicates that the extent to which CR-associated survival effects are explained by TMB is cancer-type dependent (**Supplementary Figure 14**).

### CR-altered and CR-unaltered tumors exhibit cancer-specific differences in the tumor microenvironment

We next examined whether CR mutations were associated with differences in tumor microenvironment (TME) composition (**Supplementary Figure 15**). Bulk RNA-seq deconvolution of TCGA samples using quanTIseq and EPIC revealed marked cancer-specific variation, with no uniform immune-cell pattern distinguishing CR-altered from CR-unaltered tumors across cancer types (**Supplementary Figure 15A and C**). RCC showed some of the most pronounced changes across multiple immune-cell populations, while CRC and ESCA/STAD also displayed substantial shifts in selected macrophage and lymphocyte populations. ESTIMATE similarly revealed cancer-specific differences, with higher immune, stromal, and composite scores particularly evident in RCC and CRC, whereas PAAD and several other cancers showed lower or mixed patterns (**Supplementary Figure 15B**). Similar cancer-specific heterogeneity was observed when the analysis was restricted to the pro-survival CR sets derived from the combined MSK-ICI cohort, with RCC, CRC, and ESCA/STAD showing the most prominent immune-cell alterations and other cancer types displaying weaker or mixed changes (**Supplementary Figure 16**). Overall, CR alterations were associated with measurable but strongly cancer-dependent changes in TME composition rather than a uniform pan-cancer immune phenotype.

### Independent validation of cancer-specific pro-survival CR genes and their association with TMB

The discovery findings were evaluated in the independent AACR Project GENIE cohort. To improve comparability, IPTW was applied both between the MSK-ICI discovery and GENIE-ICI cohorts and between GENIE ICI and GENIE non-ICI cohorts, balancing age, sex, and metastatic status where available before survival analyses (**Supplementary Figure 17**). The discovery-defined CR gene sets were then applied to GENIE without further gene selection. For the full set of 26 CRs and cancer-specific pro-survival CR sets, survival associations were directionally concordant between MSK-ICI and GENIE-ICI in most of the evaluated cancer types, although GENIE estimates were generally weaker and less precise, consistent with its smaller cancer-specific ICI subsets (**Figure 4A and B**). CRC showed the clearest cross-cohort reproducibility, while PRAD showed discordant directionality. Within GENIE, co-occurrence of ≥2 recurrent CR mutations was significantly associated with improved survival in ICI-treated patients compared with either no recurrent CR mutations (HR=0.67, 95% CI 0.50–0.91) or a single mutation (HR=0.65, 95% CI 0.47– 0.89), whereas the corresponding non-ICI comparisons were null (**Figure 4C; Supplementary Figure 18A**). Notably, co-occurrence across the complete 26-CR set followed the same directional trend toward improved survival in GENIE-ICI, although the association was weaker and did not reach statistical significance (**Supplementary Figure 18B**).

**Figure 4.**
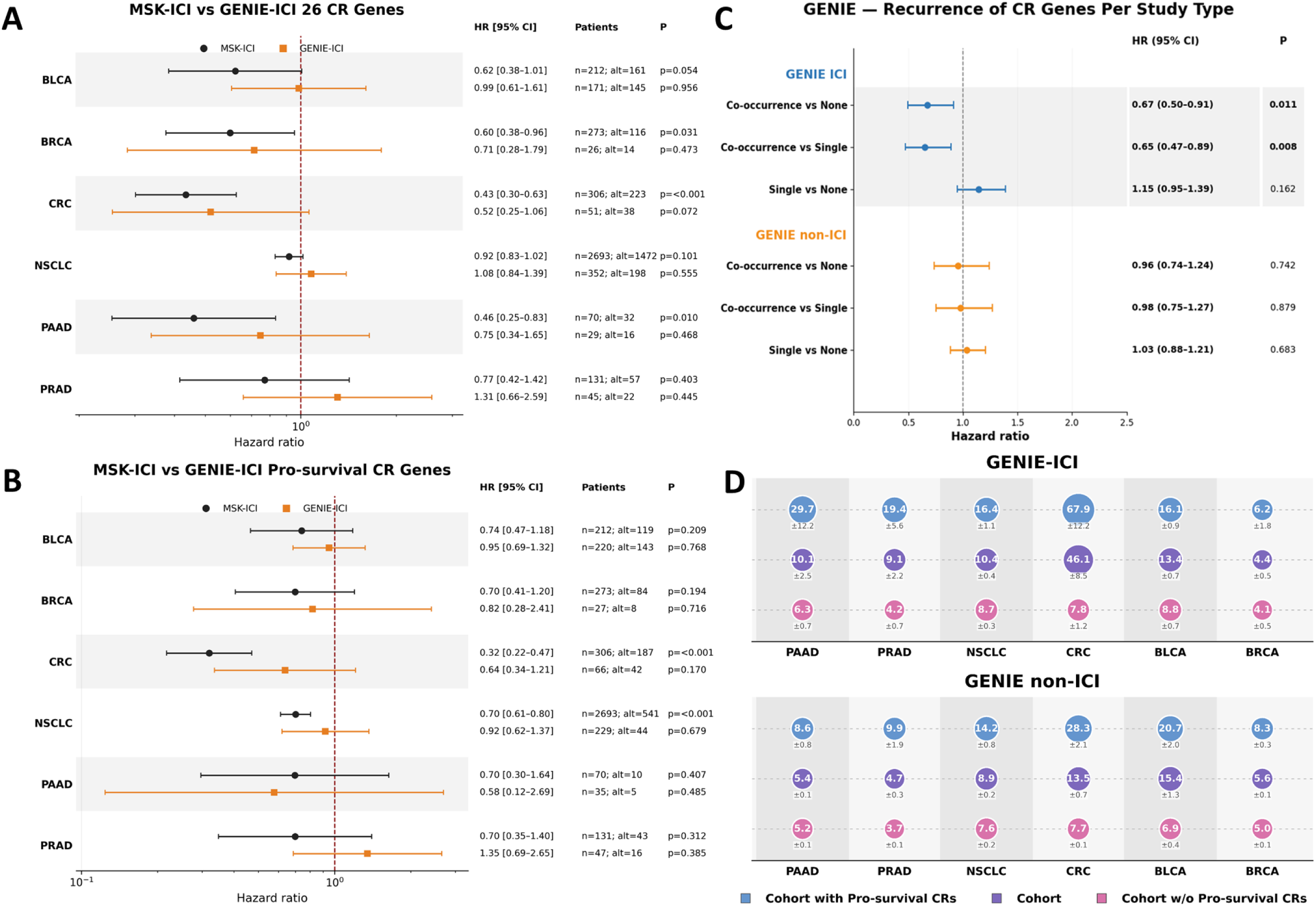
Independent validation of CR-associated survival patterns and TMB in AACR Project GENIE IPTW-weighted cohort. (A) Forest plot comparing survival associations of the complete 26-CR gene between the IPTW-weighted MSK-ICI discovery cohort and the independent GENIE-ICI cohort across six cancer types. (B) Corresponding comparison for tumors altered in the cancer-specific pro-survival CR gene sets. Points represent hazard ratios (HRs) and horizontal lines 95% confidence intervals (CIs); patient numbers, numbers of CR-altered cases, and p-values are shown. (C) Survival associations of recurrent CR mutation co-occurrence in the IPTW-weighted GENIE ICI and GENIE non-ICI cohorts, comparing tumors with ≥2 recurrent CR mutations with tumors harboring either a single or no recurrent CR mutation. (D) Tumor mutational burden (TMB) in GENIE ICI and GENIE non-ICI cohorts for tumors harboring cancer-specific pro-survival CR alterations, the complete cancer cohort, and tumors lacking these alterations. Circle size is proportional to TMB, with the corresponding TMB value shown within each circle and variability indicated below. Colors denote the respective cohorts and CR-defined groups as indicated. This figure is related to Supplementary Figures 17-19. The n values for all analyzed groups can be found in Supplementary Data.

The association between CR alterations and increased TMB was also reproduced in GENIE. Tumors altered in the complete 26-CR set as well as in the cancer-specific pro-survival CR alterations showed significantly higher TMB than CR-unaltered tumors across all six cancer types in both ICI and non-ICI cohorts (**Figure 4D; Supplementary Figure 19A and B**). TMB also generally increased with the number of recurrent CR alterations, with tumors harboring ≥2 recurrent mutations showing the highest TMB in all cancer types (**Supplementary Figure 19C**). Together, these independent analyses support the cross-cohort reproducibility of the CR-associated survival signal, particularly for recurrent CR co-occurrence, and confirm the consistent relationship between CR alterations and elevated TMB.

## DISCUSSION

Our findings reveal a structured relationship between chromatin-regulator mutations and survival following immune checkpoint therapy. Moving beyond our previous pan-cancer observations, we examined these associations across discovery and independent validation cohorts comprising thousands of ICI-treated and non-ICI-treated patients, revealing several distinct layers of association. First, we identified cancer-specific CR mutation patterns associated with favorable survival and then found that a subset of these genes recurred across multiple cancer types. Co-occurrence of CR alterations was associated with a further survival advantage in ICI-treated cohorts and with progressively increased TMB, while individual CR genes showed particularly robust associations in selected cancers. Finally, bulk RNA-seq analyses indicated that the TME consequences of CR alterations are strongly cancer-dependent rather than uniform across tumors. Together, these results suggest that CR alterations comprise both cancer-specific and shared components relevant to ICI-associated outcomes.

Our results are consistent with a growing literature linking mutations in individual chromatin regulators to favorable outcomes following ICI treatment. Alterations in SWI/SNF components, particularly *ARID1A*, *ARID1B*, and *ARID2*, have been associated with improved ICI outcomes across multiple cancer types (20–24). Similarly, *SETD2* mutations have been linked to increased TMB and MSI, altered immune-related transcriptional programs, and favorable outcomes following ICI therapy (25,26). *KMT2C* and *KMT2D* alterations have also been associated with ICI benefit in colorectal cancer and NSCLC (27–29), while *TET1* mutations have been associated with increased TMB, enhanced immune activity, and improved checkpoint-blockade outcomes (30–32), suggesting that this phenomenon extends across distinct classes of epigenetic regulators. Consistent with these studies, several of these genes also emerged among the recurrent or cancer-specific CRs identified in our analysis, supporting convergence between our findings and previous reports.

Tumor microenvironment analyses further indicated that the consequences of CR alterations are strongly cancer-dependent. Transcriptomic deconvolution revealed no uniform immune phenotype, with immune and stromal changes varying across cancer types and CR gene sets and being most pronounced in RCC, CRC, and ESCA/STAD. These findings suggest that CR dysfunction may interact with lineage-specific transcriptional and epigenetic programs to shape distinct tumor microenvironments.

CR mutations may promote ICI responsiveness through at least two non-exclusive mechanisms: epigenetic dysregulation could impair genome maintenance and increase mutational/neoantigen burden, although our observations showing CR-associated ICI benefit after adjustment for TMB indicate that TMB is unlikely to provide a complete explanation. Another possible mechanism is that altered chromatin accessibility may increase the expression or presentation of existing neoantigens and, more broadly, reshape the tumor immune environment through differential expression of immune-related genes. These possibilities warrant direct functional investigation, highlighting the central role of epigenetic dysregulation in cancer development, progression, and therapeutic response.

The study has several limitations that should be taken into account when interpreting the findings. Because the analysis was retrospective and based on publicly available clinicogenomic datasets, treatment allocation was not randomized. IPTW improved covariate balance, but some residual imbalance persisted, most notably in MSK-ICI versus TCGA comparisons. This is particularly relevant because TCGA is composed predominantly of primary tumors, whereas MSK-ICI largely includes patients with advanced disease. TCGA should therefore be regarded as a heterogeneous non-ICI comparison cohort rather than a true untreated control. Analyses performed against the MSK-CHORD non-ICI reduce some of these cross-cohort differences, although treatment-selection bias and unmeasured confounding remain possible.

There are also technical and endpoint-related limitations. Differences between whole-exome sequencing and targeted panel sequencing may affect mutation detection and cross-cohort comparability. TME deconvolution has several methodological limitations; EPIC and quanTIseq rely on fixed reference signatures that may not fully capture cancer-specific immune and stromal states, while ESTIMATE provides relative scores rather than absolute cell fractions. Differences in reference panels and cell-type definitions can also lead to partial discordance between methods. Moreover, in cancers with high baseline mortality, such as NSCLC, overall survival may be relatively insensitive to treatment-associated differences, and future studies may benefit from progression-free survival or direct response measures. Finally, the small number of CR-altered cases in several cancer types reduced statistical power and limited the robustness of some cancer-specific conclusions.

In conclusion, our findings identify cancer-specific and recurrent CR mutation patterns associated with improved outcomes following ICI therapy, with co-occurring CR alterations and increased TMB further strengthening this association. These results highlight genetically driven epigenetic dysregulation as an important feature of cancer biology and a potential determinant of response to immune checkpoint therapy.

## Declarations

### Ethics approval and consent to participate

Not applicable. This study used exclusively publicly available, de-identified data obtained from previously published cancer genomics datasets and projects; therefore, no additional ethics approval or informed consent was required.

### Consent for publication

Not applicable. This study used only publicly available, de-identified data that have been previously reported in the scientific literature.

### Conflict of interest

The authors declare that the research was conducted in the absence of any commercial or financial relationships that could be perceived as potential conflicts of interest.

### Availability of data and material

All data analyzed in this study are publicly available through the cBioPortal for Cancer Genomics, http://www.cbioportal.org and https://genie.cbioportal.org.

### Generative AI statement

GPT-5.6 Sol (OpenAI) and Grammarly were utilized for language editing and stylistic improvement. No AI tools were used for data analysis, data interpretation, or the generation of scientific conclusions. All content was reviewed and approved by the authors.

Any alternative text (alt text) provided alongside figures in this article has been generated by Frontiers with the support of artificial intelligence, and reasonable efforts have been made to ensure accuracy, including review by the authors wherever possible. If you identify any issues, please contact us.

## Funding

No dedicated research funding was received. Personnel salaries were supported by Fingerprint Diagnostics LLC and Zan Mitrev Clinic, while all analyzed data were publicly available.

### Author contributions

GK conceived and designed the study. DjB, MG, SK, MN, and LM collected, organized, curated, and analyzed the data. DjB performed bioinformatic analyses. IK and ZM contributed scientific input and interpretation. GK drafted the manuscript. All authors contributed to manuscript revision, reviewed the final version, and approved it for publication.

## Notes

### Competing Interest Statement

The authors have declared no competing interest.

