## Supplementary Figures for "Cancer type-specific profiling of chromatin regulator mutations identifies recurrent and co-occurring gene sets associated with improved survival following immune checkpoint therapy"

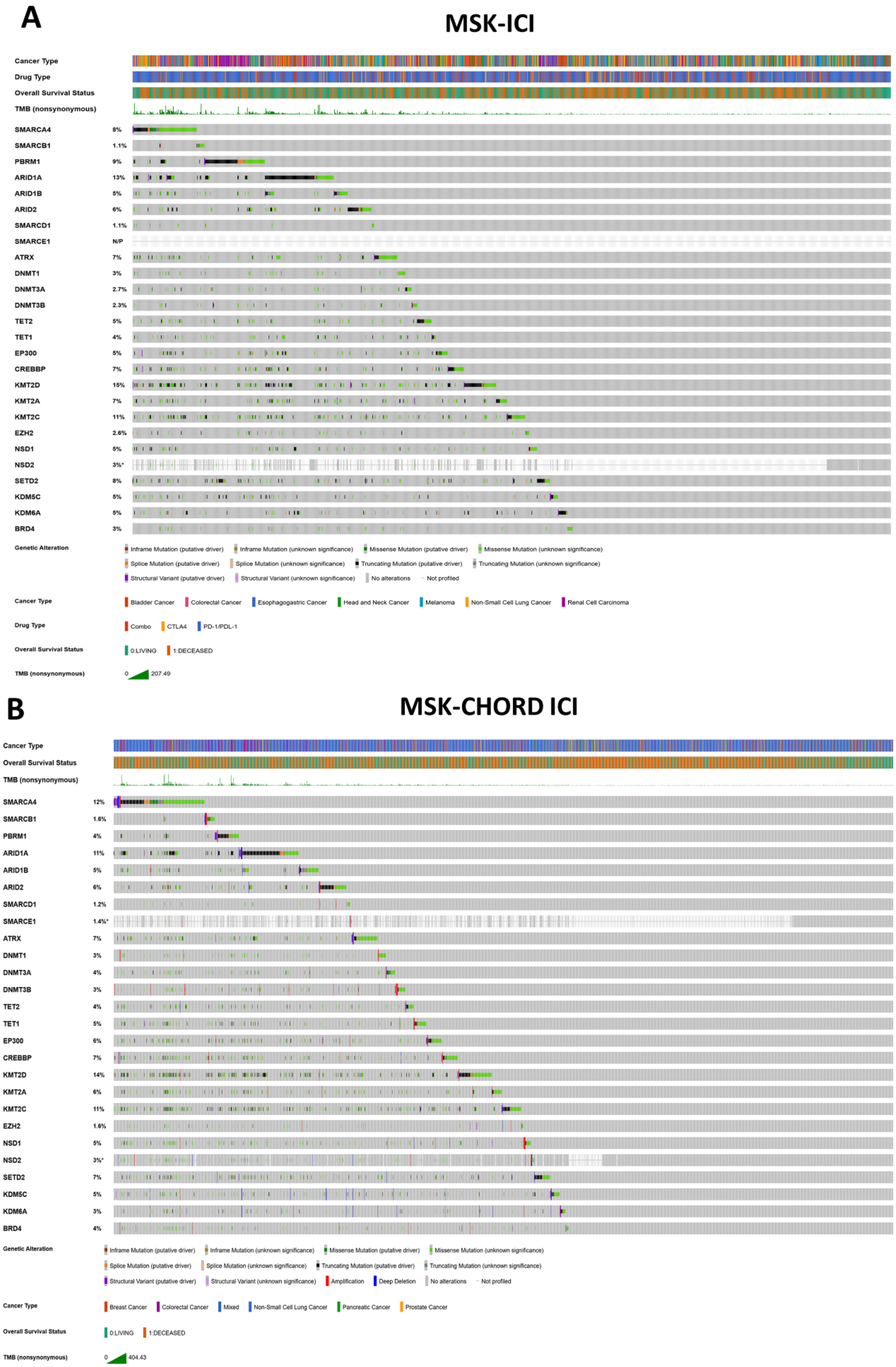
A
MSK-ICI
Cancer Type
Drug Type
Overall Survival Status
TMB (nonsynonymous)
SMARCA4 8%
SMARCB1 1.1%
PBRM1 9%
ARID1A 13%
ARID1B 5%
ARID2 6%
SMARCD1 1.1%
SMARCE1 N/P
ATRX 7%
DNMT1 3%
DNMT3A 2.7%
DNMT3B 2.3%
TET2 5%
TET1 4%
EP300 5%
CREBBP 7%
KMT2D 15%
KMT2A 7%
KMT2C 11%
EZH2 2.6%
NSD1 5%
NSD2 3%*
SETD2 8%
KDM5C 5%
KDM6A 5%
BRD4 3%
Genetic Alteration
Inframe Mutation (putative driver)
Inframe Mutation (unknown significance)
Missense Mutation (putative driver)
Missense Mutation (unknown significance)
Splice Mutation (putative driver)
Splice Mutation (unknown significance)
Truncating Mutation (putative driver)
Truncating Mutation (unknown significance)
Structural Variant (putative driver)
Structural Variant (unknown significance)
No alterations
Not profiled
Cancer Type
Bladder Cancer
Colorectal Cancer
Esophagogastric Cancer
Head and Neck Cancer
Melanoma
Non-Small Cell Lung Cancer
Renal Cell Carcinoma
Drug Type
Combo
CTLA4
PD-1/PDL-1
Overall Survival Status
0:LIVING
1:DECEASED
TMB (nonsynonymous)
0
207.49
B
MSK-CHORD ICI
Cancer Type
Overall Survival Status
TMB (nonsynonymous)
SMARCA4 12%
SMARCB1 1.6%
PBRM1 4%
ARID1A 11%
ARID1B 5%
ARID2 6%
SMARCD1 1.2%
SMARCE1 1.4%*
ATRX 7%
DNMT1 3%
DNMT3A 4%
DNMT3B 3%
TET2 4%
TET1 5%
EP300 6%
CREBBP 7%
KMT2D 14%
KMT2A 6%
KMT2C 11%
EZH2 1.6%
NSD1 5%
NSD2 3%*
SETD2 7%
KDM5C 5%
KDM6A 3%
BRD4 4%
Genetic Alteration
Inframe Mutation (putative driver)
Inframe Mutation (unknown significance)
Missense Mutation (putative driver)
Missense Mutation (unknown significance)
Splice Mutation (putative driver)
Splice Mutation (unknown significance)
Truncating Mutation (putative driver)
Truncating Mutation (unknown significance)
Structural Variant (putative driver)
Structural Variant (unknown significance)
Amplification
Deep Deletion
No alterations
Not profiled
Cancer Type
Breast Cancer
Colorectal Cancer
Mixed
Non-Small Cell Lung Cancer
Pancreatic Cancer
Prostate Cancer
Overall Survival Status
0:LIVING
1:DECEASED
TMB (nonsynonymous)
0
404.43

**Supplementary Figure 1. Genomic landscape of chromatin regulator alterations in ICI-treated cohorts.** OncoPrints show alterations across the 26 analyzed chromatin regulator (CR) genes in (A) the original MSK-ICI cohort and (B) the MSK-CHORD-ICI cohort. Columns represent individual patients and rows represent CR genes, with the percentage of altered tumors indicated for each gene. Clinical annotations include cancer type, overall survival status, and tumor mutational burden (TMB); ICI drug class is additionally shown for MSK-ICI. Colors indicate mutation or structural-variant type as defined in the legend, while gray and white denote no detected alteration and regions not profiled, respectively.

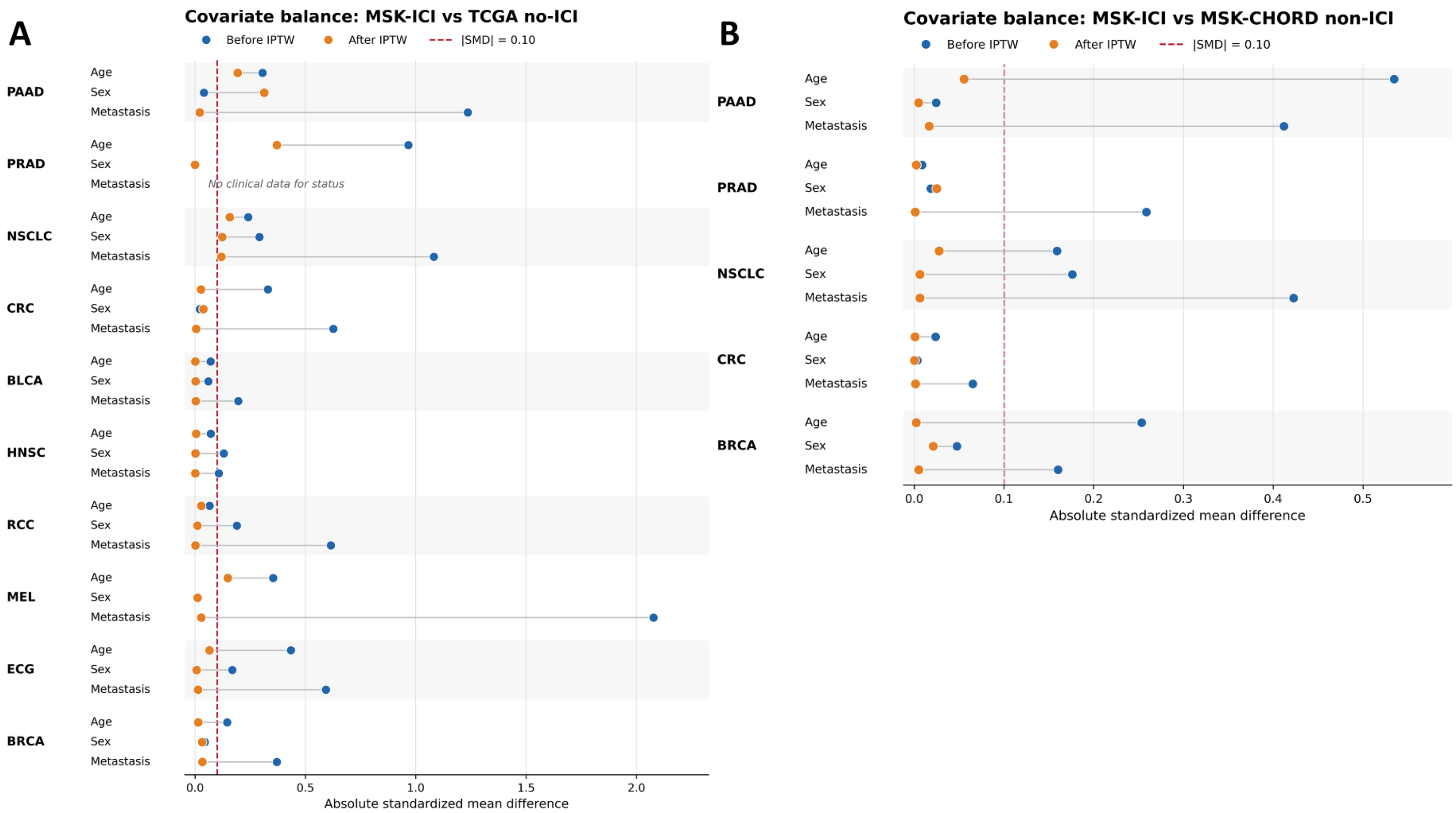


**Supplementary Figure 2. Covariate balance before and after inverse probability of treatment weighting (IPTW).** Absolute standardized mean differences (SMDs) are shown in the love plots for available baseline covariates across the two cohort comparisons. (A) MSK-ICI versus TCGA non-ICI, (B) MSK-ICI vs MSK-CHORD non-ICI. Covariates included age, sex, and metastatic status, where available. Blue points represent SMDs before IPTW and orange points represent SMDs after weighting. The dashed vertical line at SMD = 0.10 indicates the predefined threshold for adequate covariate balance; values below 0.10 were considered balanced. IPTW substantially reduced baseline imbalances across most cancer-specific comparisons, although residual imbalance remained in selected covariates and cancer types.

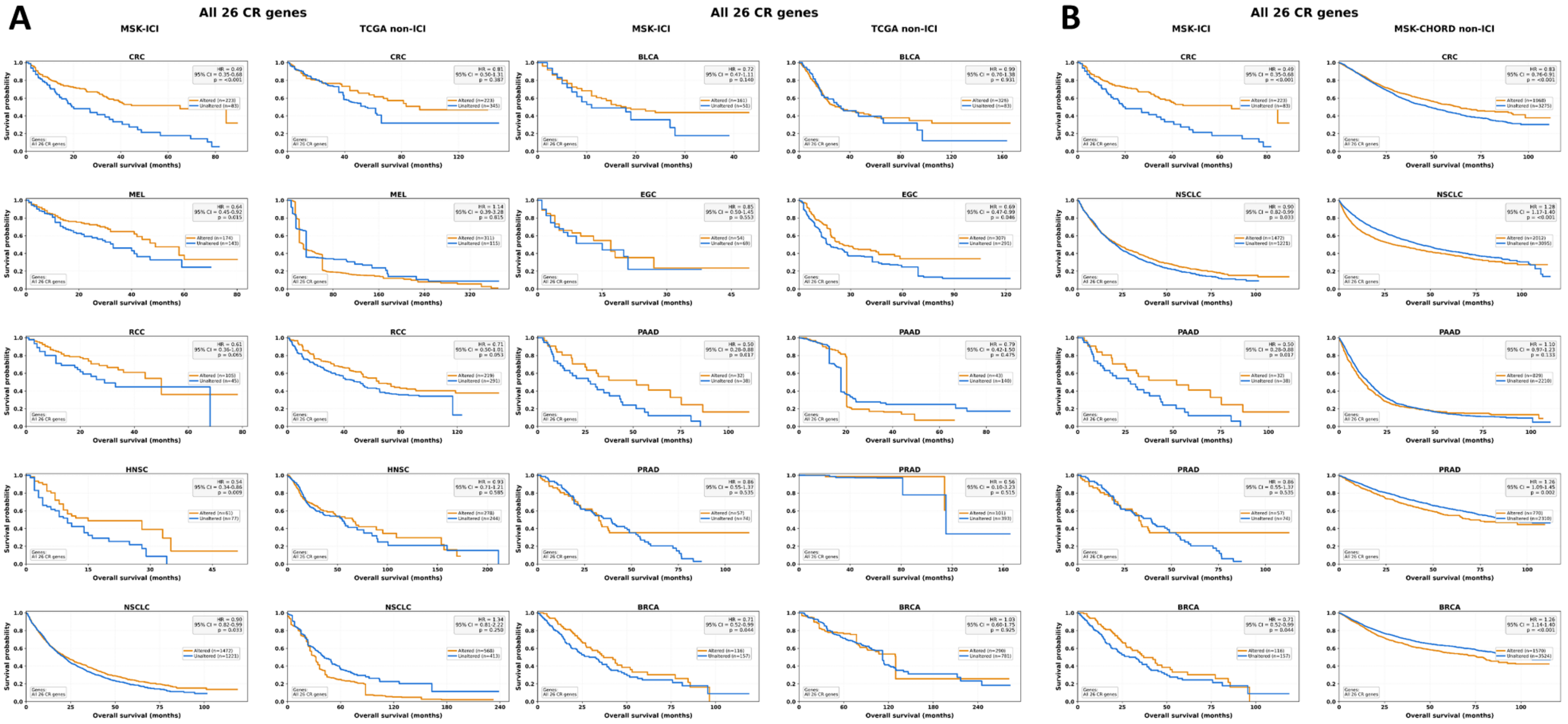


**Supplementary Figure 3. Cancer-specific overall survival associated with alterations in the complete 26-CR gene set across ICI-treated and non-ICI-treated cohorts**. IPTW-weighted Kaplan–Meier curves compare tumors harboring mutations in at least one of the 26 analyzed chromatin regulator (CR) genes with CR-unaltered tumors. (A) Comparisons between the combined MSK-ICI cohort and corresponding TCGA non-ICI cohorts across 10 cancer types. (B) Comparisons between MSK-ICI and MSK-CHORD non-ICI cohorts across the five cancer types represented in both datasets. Hazard ratios (HRs), 95% confidence intervals (CIs), and P values are shown within each panel. Orange curves indicate CR-altered tumors and blue curves CR-unaltered tumors. Overall survival (OS) is shown in months. Cancer types included colorectal cancer (CRC), melanoma (MEL), renal cell carcinoma (RCC), head and neck squamous cell carcinoma (HNSC), non-small cell lung cancer (NSCLC), bladder cancer (BLCA), esophagogastric cancer (EGC), pancreatic adenocarcinoma (PAAD), prostate adenocarcinoma (PRAD), and breast cancer (BRCA).

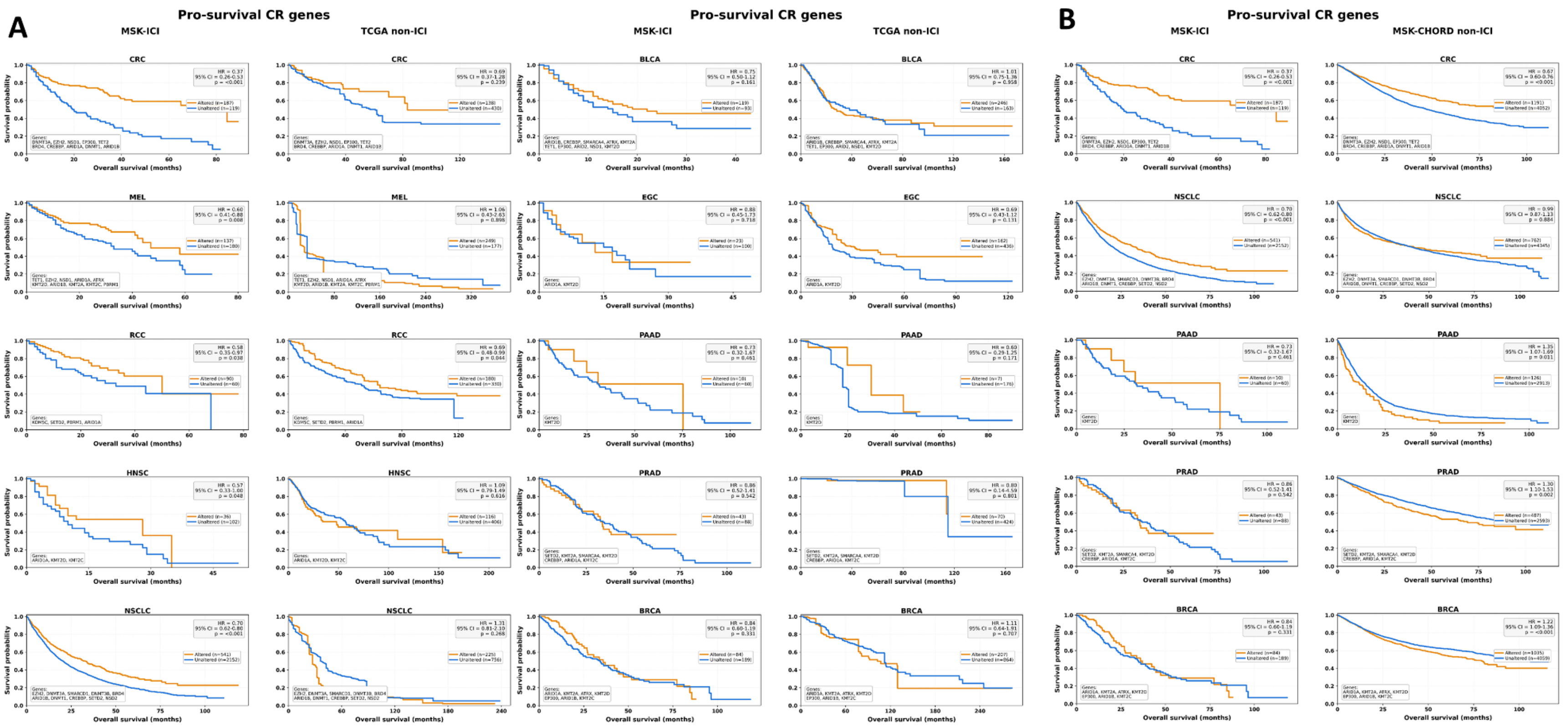


**Supplementary Figure 4. Overall survival according to cancer-specific pro-survival chromatin regulator mutations in MSK-ICI and matched TCGA non-ICI cohorts.** . IPTW-weighted Kaplan–Meier curves compare tumors harboring mutations in at least one of cancer-specific pro-survival chromatin regulators with CR-unaltered tumors. (A) Comparisons between the combined MSK-ICI cohort and corresponding TCGA non-ICI cohorts across 10 cancer types. (B) Comparisons between MSK-ICI and MSK-CHORD non-ICI cohorts across the five cancer types represented in both datasets. Hazard ratios (HRs), 95% confidence intervals (CIs), and P values are shown within each panel. Orange curves indicate CR-altered tumors and blue curves CR-unaltered tumors. Overall survival (OS) is shown in months. Cancer types included colorectal cancer (CRC), melanoma (MEL), renal cell carcinoma (RCC), head and neck squamous cell carcinoma (HNSC), non-small cell lung cancer (NSCLC), bladder cancer (BLCA), esophagogastric cancer (EGC), pancreatic adenocarcinoma (PAAD), prostate adenocarcinoma (PRAD), and breast cancer (BRCA).

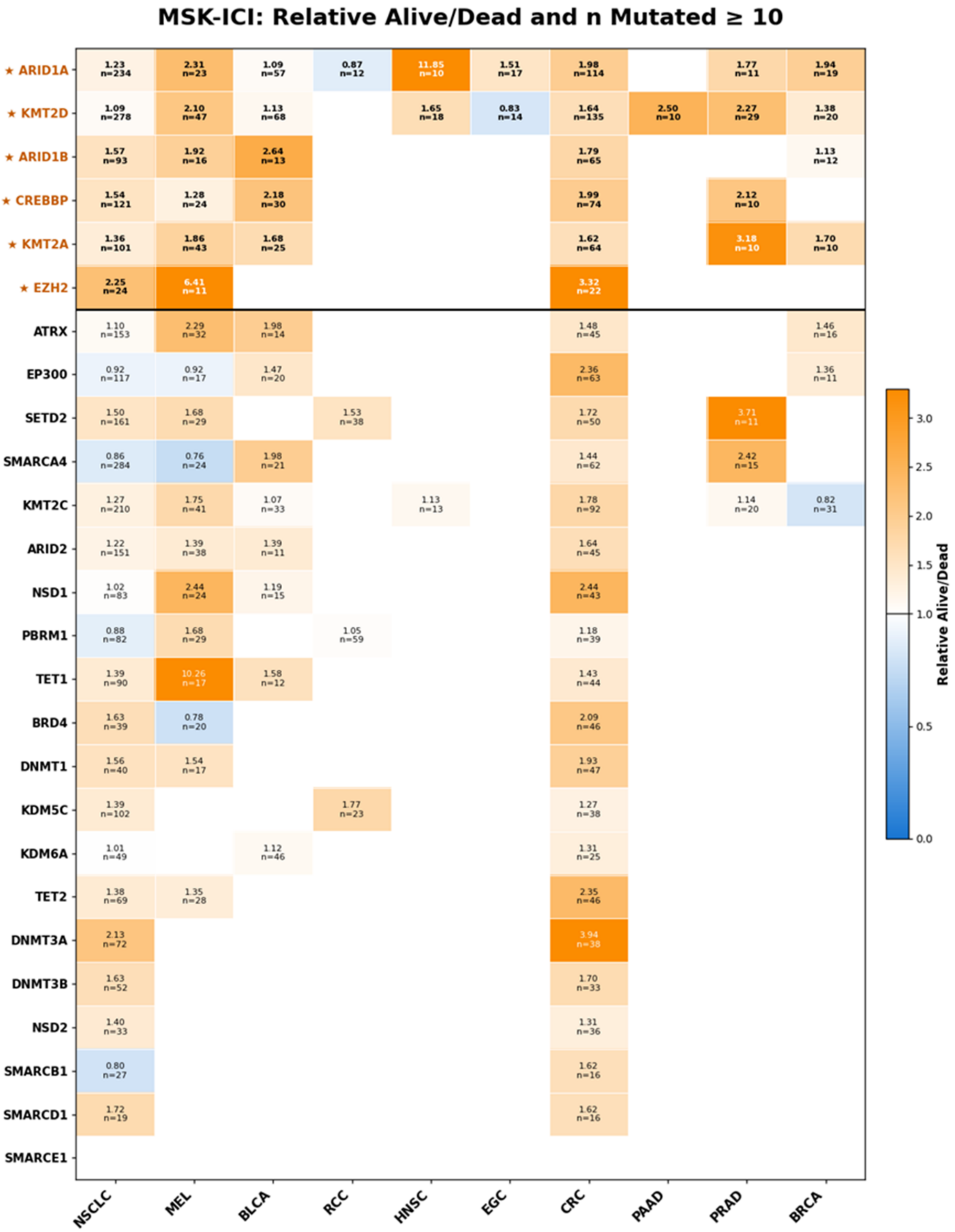
MSK-ICI: Relative Alive/Dead and n Mutated ≥ 10
★ ARID1A
1.23 n=234
2.31 n=23
1.09 n=57
0.87 n=12
11.85 n=10
1.51 n=17
1.98 n=114
1.77 n=11
1.94 n=19
★ KMT2D
1.09 n=278
2.10 n=47
1.13 n=68
1.65 n=18
0.83 n=14
1.64 n=135
2.50 n=10
2.27 n=29
1.38 n=20
★ ARID1B
1.57 n=93
1.92 n=16
2.64 n=13
1.79 n=65
1.13 n=12
★ CREBBP
1.54 n=121
1.28 n=24
2.18 n=30
1.99 n=74
2.12 n=10
★ KMT2A
1.36 n=101
1.86 n=43
1.68 n=25
1.62 n=64
3.18 n=10
1.70 n=10
★ EZH2
2.25 n=24
6.41 n=11
3.32 n=22
ATRX
1.10 n=153
2.29 n=32
1.98 n=14
1.48 n=45
1.46 n=16
EP300
0.92 n=117
0.92 n=17
1.47 n=20
2.36 n=63
1.36 n=11
SETD2
1.50 n=161
1.68 n=29
1.53 n=38
1.72 n=50
3.71 n=11
SMARCA4
0.86 n=284
0.76 n=24
1.98 n=21
1.44 n=62
2.42 n=15
KMT2C
1.27 n=210
1.75 n=41
1.07 n=33
1.13 n=13
1.78 n=92
1.14 n=20
0.82 n=31
ARID2
1.22 n=151
1.39 n=38
1.39 n=11
1.64 n=45
NSD1
1.02 n=83
2.44 n=24
1.19 n=15
2.44 n=43
PBRM1
0.88 n=82
1.68 n=29
1.05 n=59
1.18 n=39
TET1
1.39 n=90
10.26 n=17
1.58 n=12
1.43 n=44
BRD4
1.63 n=39
0.78 n=20
2.09 n=46
DNMT1
1.56 n=40
1.54 n=17
1.93 n=47
KDM5C
1.39 n=102
1.77 n=23
1.27 n=38
KDM6A
1.01 n=49
1.12 n=46
1.31 n=25
TET2
1.38 n=69
1.35 n=28
2.35 n=46
DNMT3A
2.13 n=72
3.94 n=38
DNMT3B
1.63 n=52
1.70 n=33
NSD2
1.40 n=33
1.31 n=36
SMARCB1
0.80 n=27
1.62 n=16
SMARCD1
1.72 n=19
1.62 n=16
SMARCE1
NSCLC
MEL
BLCA
RCC
HNSC
EGC
CRC
PAAD
PRAD
BRCA
Relative Alive/Dead
3.0
2.5
2.0
1.5
1.0
0.5
0.0

55

**Supplementary Figure 5. Cancer-specific ranking of alive/dead ratios for mutated chromatin regulator genes in the MSK-ICI cohort**. Heatmap shows the relative alive/dead ratio among mutation carriers for each of the 26 analyzed chromatin regulator (CR) genes across cancer types, restricted to gene–cancer combinations with ≥10 mutated patients. Values within cells indicate the relative alive/dead ratio and the number of mutation carriers (n); blank cells indicate fewer than 10 mutated cases. Blue denotes lower and orange higher relative alive/dead ratios. The six prioritized recurrent CR genes—*ARID1A, KMT2D, ARID1B, CREBBP, KMT2A,* and *EZH2*—are marked with stars and separated from the remaining genes by the horizontal line. Cancer types include non-small cell lung cancer (NSCLC), melanoma (MEL), bladder cancer (BLCA), renal cell carcinoma (RCC), head and neck squamous cell carcinoma (HNSC), esophagogastric cancer (EGC), colorectal cancer (CRC), pancreatic adenocarcinoma (PAAD), prostate adenocarcinoma (PRAD), and breast cancer (BRCA).

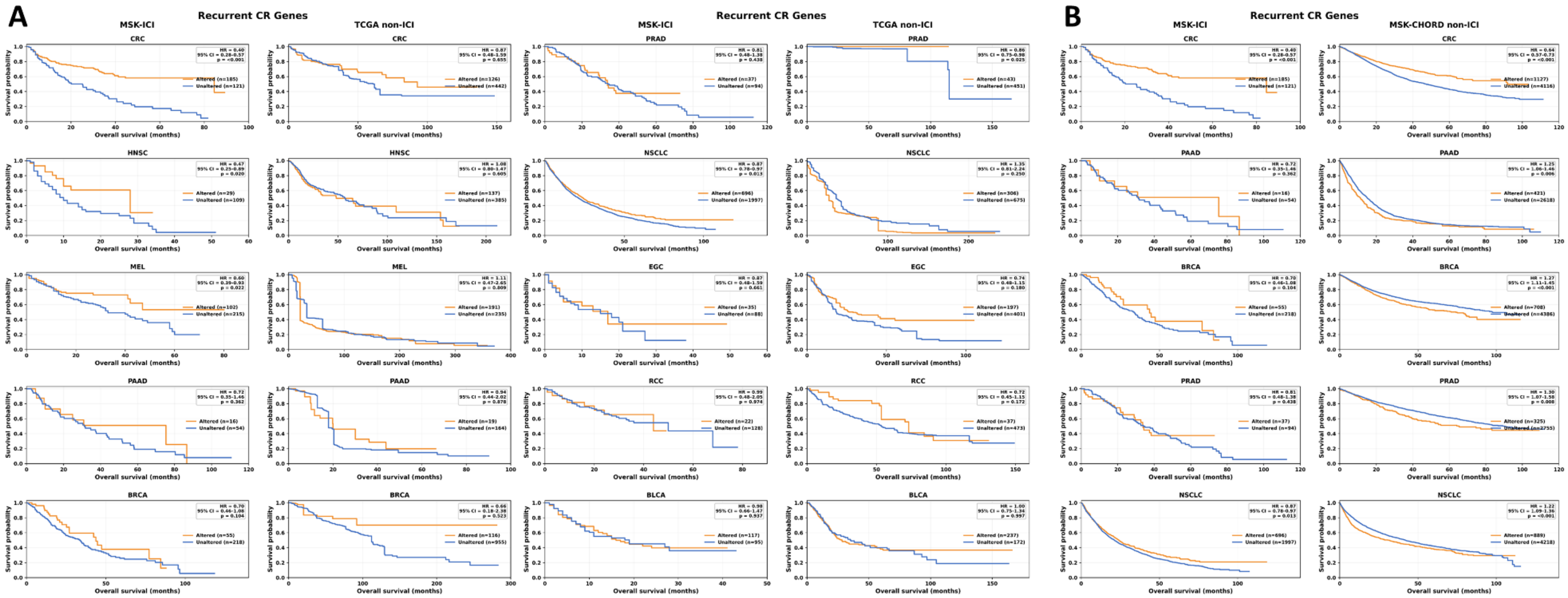


**Supplementary Figure 6. Cancer-specific overall survival associated with recurrent chromatin regulator alterations in ICI-treated and non-ICI-treated cohorts**. IPTW-weighted Kaplan–Meier curves compare tumors harboring mutations in at least one of the six recurrent CR genes (*ARID1A, KMT2D, ARID1B, CREBBP, KMT2A,* and *EZH2*) with tumors lacking these alterations. (A) Comparisons between the combined MSK-ICI cohort and corresponding TCGA non-ICI cohorts across colorectal cancer (CRC), head and neck squamous cell carcinoma (HNSC), melanoma (MEL), pancreatic adenocarcinoma (PAAD), breast cancer (BRCA), prostate adenocarcinoma (PRAD), non-small cell lung cancer (NSCLC), esophagogastric cancer (EGC), renal cell carcinoma (RCC), and bladder cancer (BLCA). (B) Comparisons between MSK-ICI and MSK-CHORD non-ICI cohorts for CRC, PAAD, BRCA, PRAD, and NSCLC. Hazard ratios (HRs), 95% confidence intervals (CIs), and P values are shown within each panel. Orange curves indicate recurrent CR-altered tumors and blue curves CR-unaltered tumors.

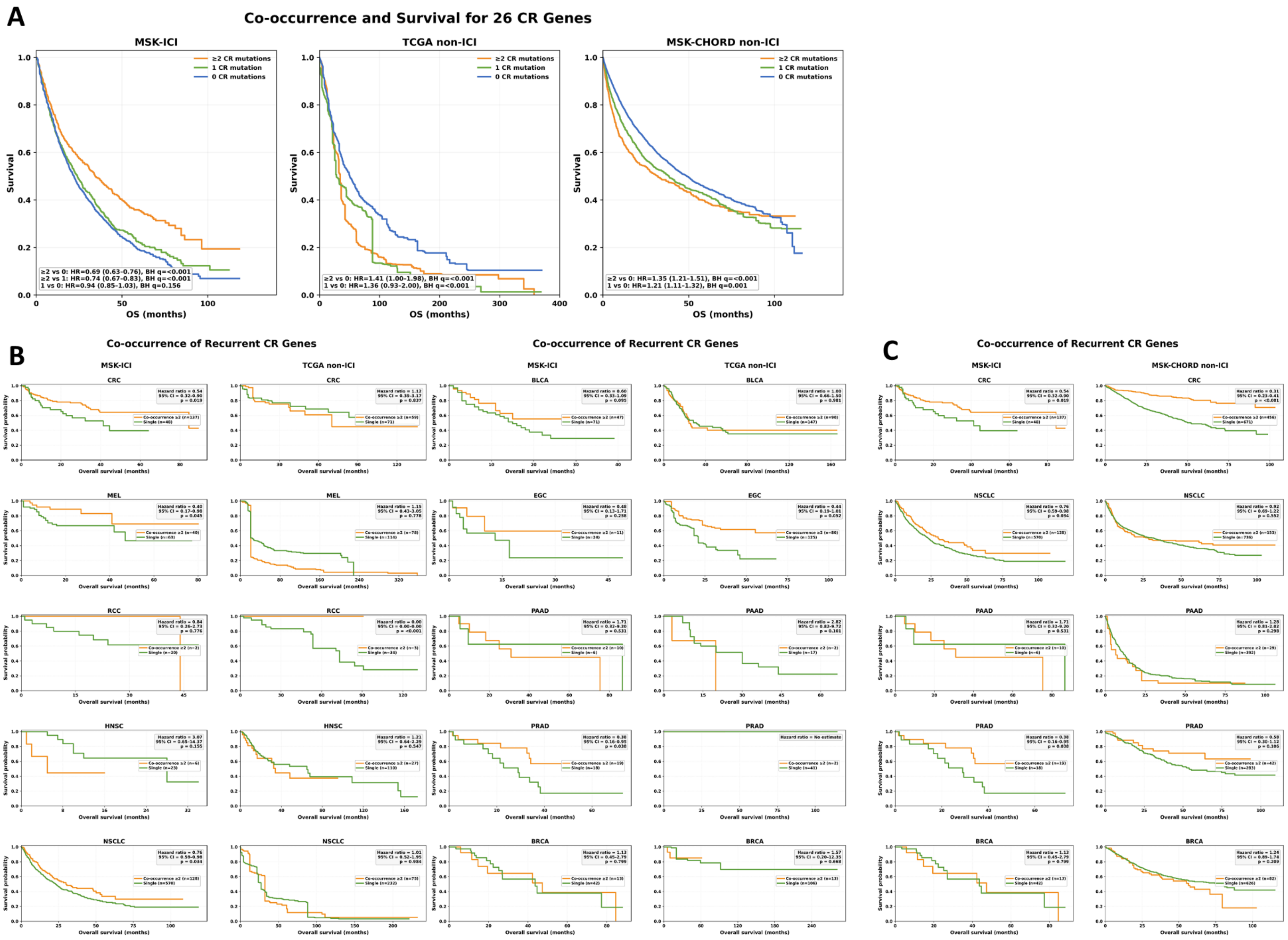
A
Co-occurrence and Survival for 26 CR Genes
MSK-ICI
TCGA non-ICI
MSK-CHORD non-ICI
≥2 CR mutations
1 CR mutation
0 CR mutations
Survival
OS (months)
≥2 vs 0: HR=0.69 (0.63-0.76), BH q=<0.001
≥2 vs 1: HR=0.74 (0.67-0.83), BH q=<0.001
1 vs 0: HR=0.94 (0.85-1.03), BH q=0.156
≥2 vs 0: HR=1.41 (1.00-1.98), BH q=<0.001
1 vs 0: HR=1.36 (0.93-2.00), BH q=<0.001
≥2 vs 0: HR=1.35 (1.21-1.51), BH q=<0.001
1 vs 0: HR=1.21 (1.11-1.32), BH q=0.001
B
Co-occurrence of Recurrent CR Genes
MSK-ICI
TCGA non-ICI
CRC
MEL
RCC
HNSC
NSCLC
BLCA
EGC
PAAD
PRAD
BRCA
C
Co-occurrence of Recurrent CR Genes
MSK-ICI
MSK-CHORD non-ICI
Survival probability
Overall survival (months)

**Supplementary Figure 7. Survival associations of co-occurring chromatin regulator mutations in ICI-treated and non-ICI-treated cohorts**. IPTW-weighted Kaplan–Meier analyses evaluate overall survival according to CR mutation burden. (A) Pan-cancer survival for the complete 26-CR gene set, comparing tumors with ≥2, 1, or 0 CR mutations in MSK-ICI, TCGA non-ICI, and MSK-CHORD non-ICI cohorts. (B–C) Cancer-specific survival analyses for the six recurrent CR genes, comparing tumors with ≥2 recurrent CR mutations (co-occurrence) with tumors harboring a single recurrent CR mutation across MSK-ICI and corresponding non-ICI cohorts. Cancer types include colorectal cancer (CRC), melanoma (MEL), renal cell carcinoma (RCC), head and neck squamous cell carcinoma (HNSC), non-small cell lung cancer (NSCLC), bladder cancer (BLCA), esophagogastric cancer (EGC), pancreatic adenocarcinoma (PAAD), prostate adenocarcinoma (PRAD), and breast cancer (BRCA). Hazard ratios (HRs), 95% confidence intervals (CIs), and P or BH-FDR-adjusted q-values are shown within the respective panels.

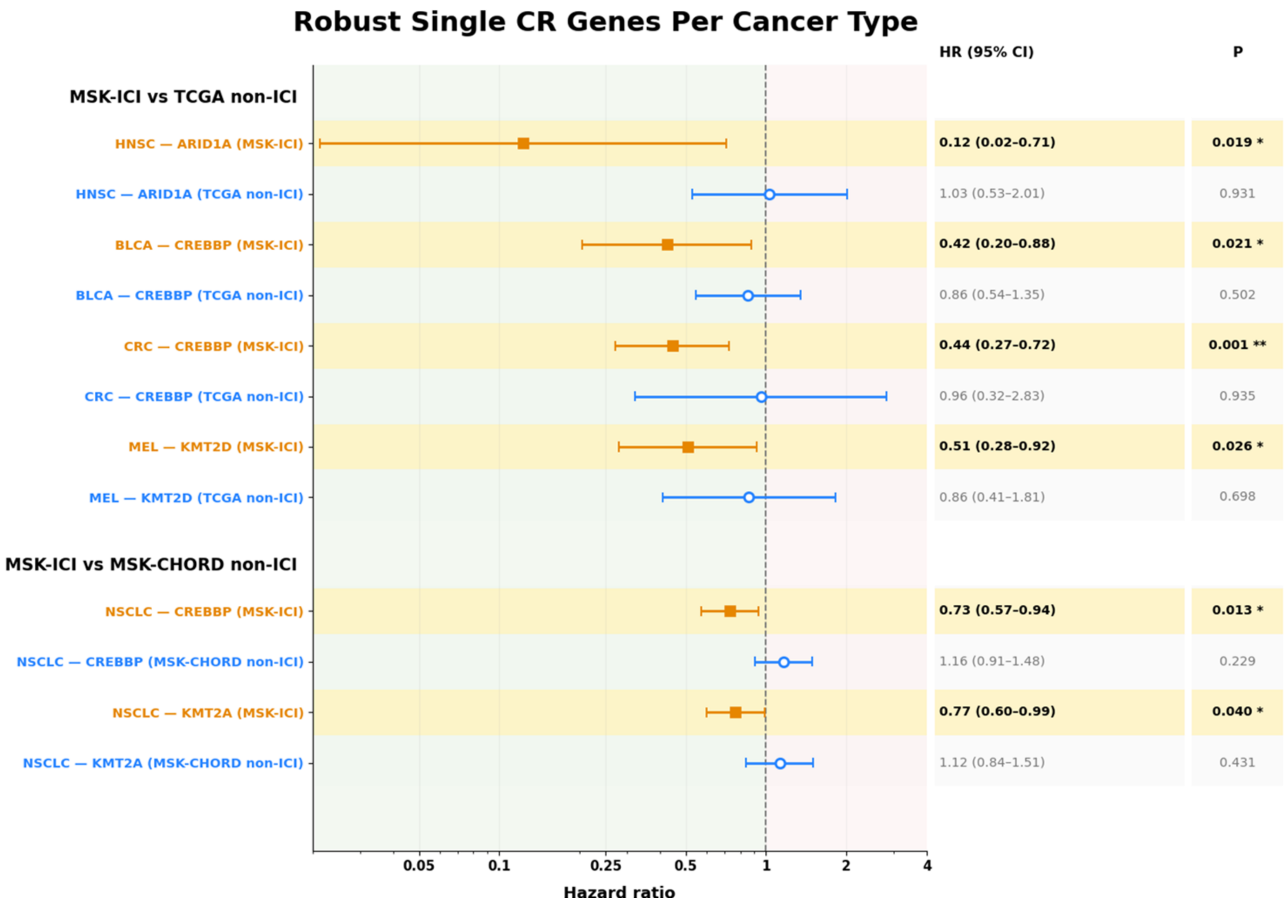


**Supplementary Figure 8. Robust cancer-specific survival associations of individual chromatin regulator genes in ICI-treated versus matched non-ICI cohorts**. Forest plot shows ATT-IPTW-weighted robust Cox estimates for individual CR gene–cancer pairs that met the predefined robustness criterion: a significant favorable association in ICI-treated patients (HR < 1, P < 0.05) without a corresponding significant favorable association in the matched non-ICI cohort. Comparisons include *ARID1A* in HNSC, CREBBP in BLCA and CRC, *KMT2D* in melanoma (MEL), and *CREBBP* and *KMT2A* in NSCLC. Orange squares indicate MSK-ICI estimates and blue circles the corresponding TCGA or MSK-CHORD non-ICI estimates. Horizontal lines represent 95% confidence intervals (CIs), and exact HRs, 95% CIs, and P values are shown on the right.

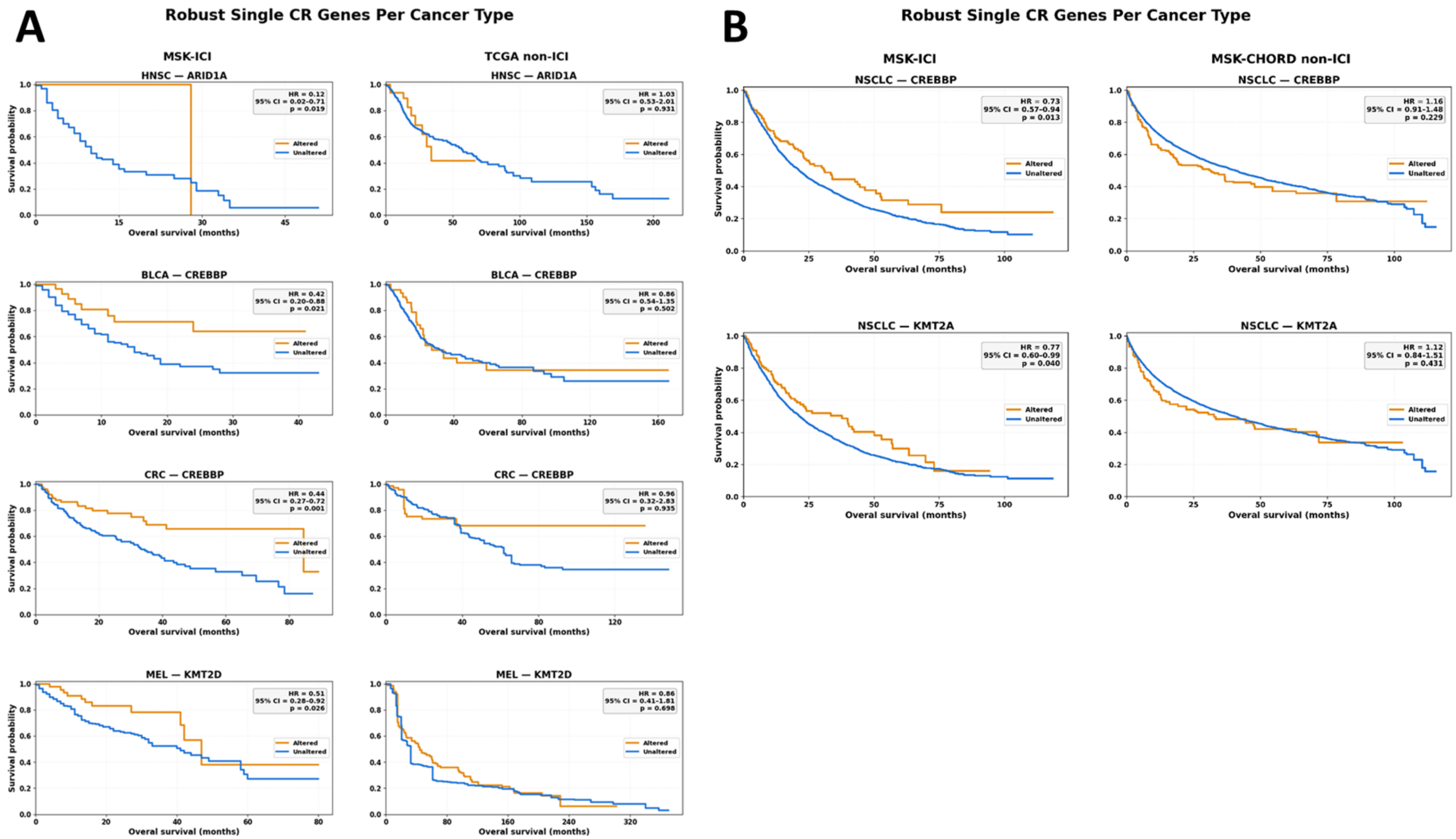


**Supplementary Figure 9. Kaplan–Meier visualization of robust cancer-specific individual chromatin regulator associations. IPTW-weighted Kaplan–**Meier curves compare overall survival between patients with mutated and unmutated tumors for the individual CR gene–cancer pairs selected by the robust Cox screening criteria. (A) Comparisons between MSK-ICI and corresponding TCGA non-ICI cohorts for *ARID1A* in HNSC, *CREBBP* in BLCA and CRC, and *KMT2D* in melanoma (MEL). (B) Comparisons between MSK-ICI and MSK-CHORD non-ICI cohorts for *CREBBP* and *KMT2A* in NSCLC. Hazard ratios (HRs), 95% confidence intervals (CIs), and P values are derived from the corresponding ATT-IPTW-weighted robust Cox models and are shown within each panel. Orange curves indicate CR-altered tumors and blue curves CR-unaltered tumors.

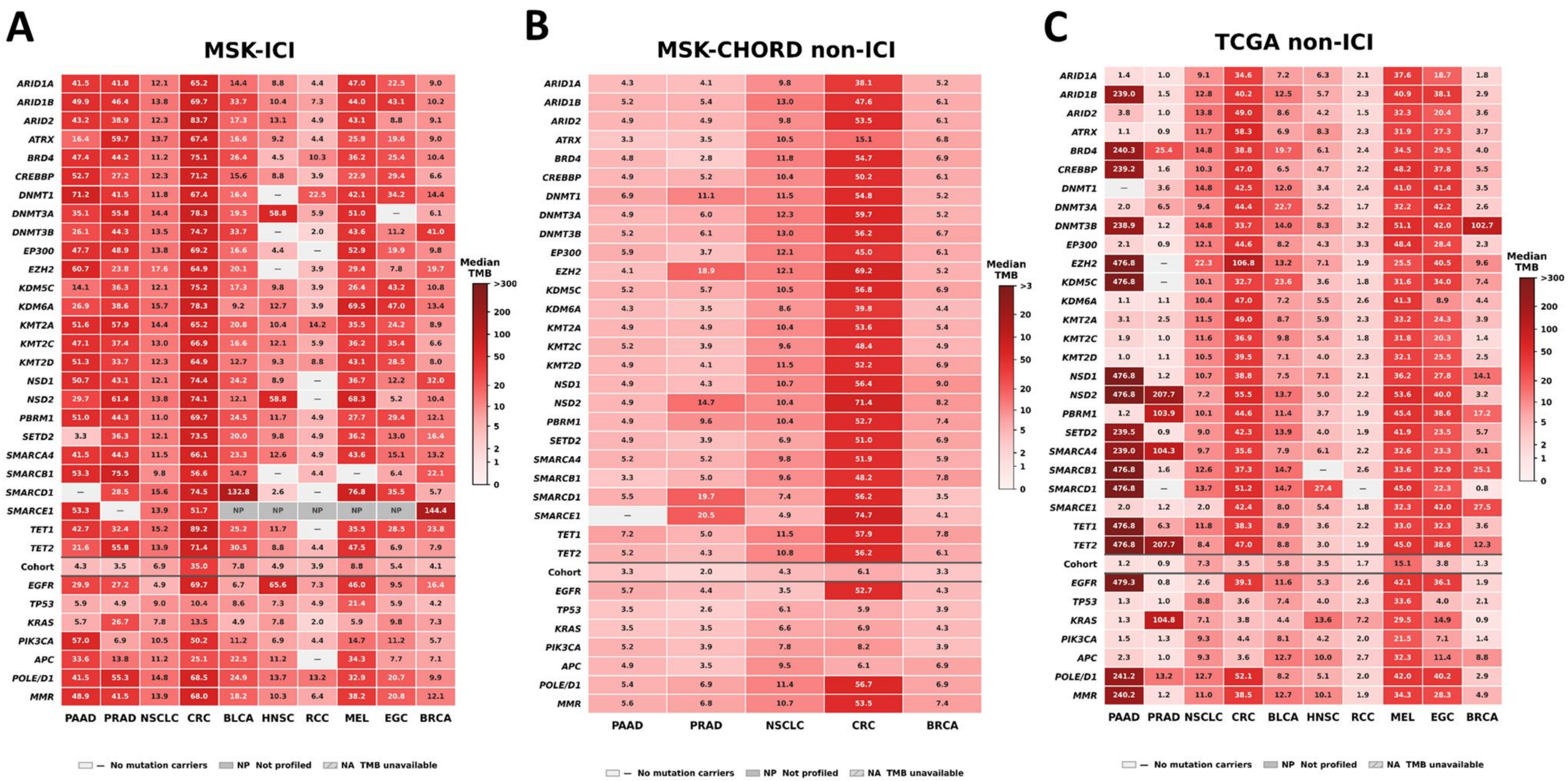


**Supplementary Figure 10. Cancer-specific tumor mutational burden associated with individual chromatin regulator mutations across ICI-treated and non-ICI-treated cohorts.** Heatmaps show median tumor mutational burden (TMB) for tumors harboring mutations in individual CR genes across cancer types in (A) MSK-ICI, (B) TCGA non-ICI and (C) MSK-CHORD non-ICI. Rows represent individual CR genes and columns represent cancer types; the corresponding overall cohort TMB and selected reference genes/pathways, including *EGFR, KRAS, TP53, PIK3CA, APC,* MMR, and *POLE/POLD1*, are shown below the dashed line. Numbers within each cell indicate the median TMB for the corresponding gene–cancer combination, with the number of mutated cases shown below. Color intensity represents relative TMB within each cancer type, with darker red indicating higher TMB. Gray cells indicate gene–

cancer combinations with no mutations among assayed samples, genes not profiled, or uncertain panel coverage, as indicated in the figure key. Cancer types include pancreatic adenocarcinoma (PAAD), prostate adenocarcinoma (PRAD), non-small cell lung cancer (NSCLC), colorectal cancer (CRC), bladder cancer (BLCA), head and neck squamous cell carcinoma (HNSC), renal cell carcinoma (RCC), melanoma (MEL), esophagogastric cancer (EGC), and breast cancer (BRCA).

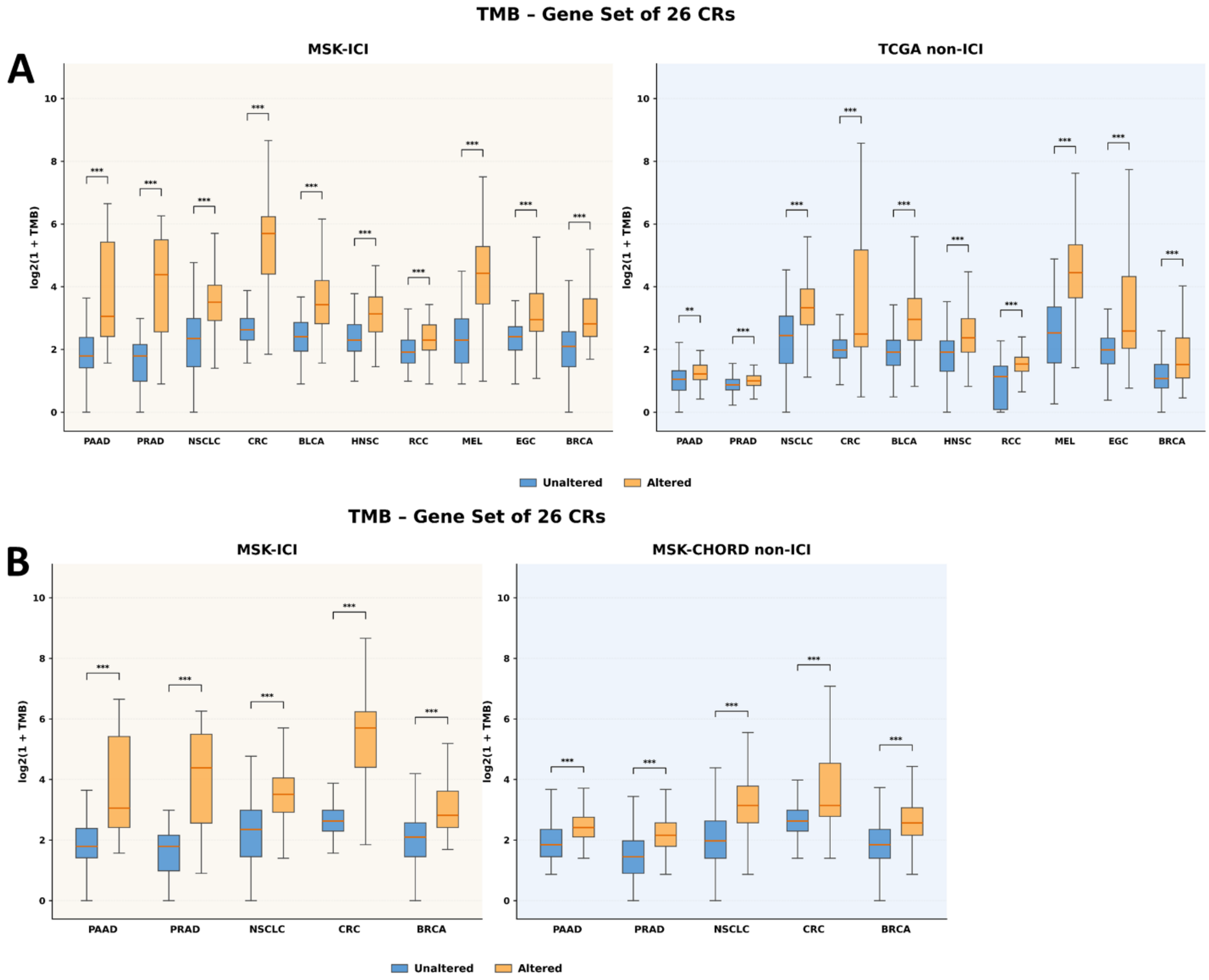


**Supplementary Figure 11. Tumor mutational burden in tumors altered and unaltered in the complete 26-CR gene set across ICI-treated and non-ICI-treated cohorts**. Boxplots show log2(1 + TMB) for tumors harboring mutations in at least one of the 26 analyzed chromatin regulator (CR) genes versus CR-unaltered tumors. (A) Comparisons between MSK-ICI and corresponding TCGA non-ICI cohorts across 10 cancer types. (B) Comparisons between MSK-ICI and MSK-CHORD non-ICI cohorts across PAAD, PRAD, NSCLC, CRC, and BRCA. Orange denotes CR-altered tumors and blue CR-unaltered tumors. Statistical significance was assessed using two-sided Mann–Whitney U tests; $**P < 0.01$ and $***P < 0.001$.

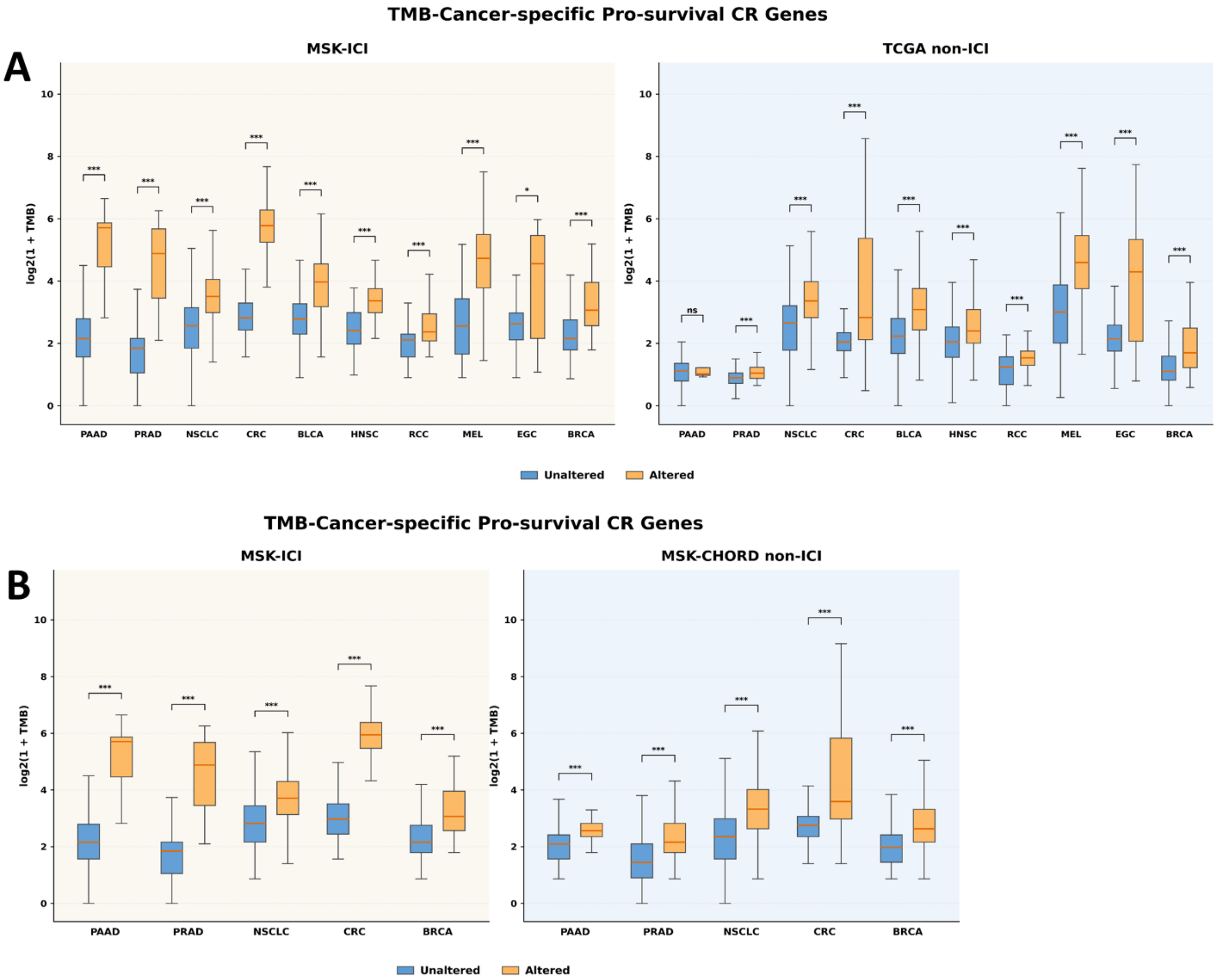


**Supplementary Figure 12. Tumor mutational burden in tumors altered and unaltered in the cancer-specific pro-survival gene sets across ICI-treated and non-ICI-treated cohorts**. Boxplots show log2(1 + TMB) for tumors harboring mutations in at least one of the cancer-specific pro-survival chromatin regulator (CR) genes versus CR-unaltered tumors. (A) Comparisons between MSK-ICI and corresponding TCGA non-ICI cohorts across 10 cancer types. (B) Comparisons between MSK-ICI and MSK-CHORD non-ICI cohorts across PAAD, PRAD, NSCLC, CRC, and BRCA. Orange denotes CR-altered tumors and blue CR-unaltered tumors. Statistical significance was assessed using two-sided Mann–Whitney U tests; **P < 0.01 and ***P < 0.001.

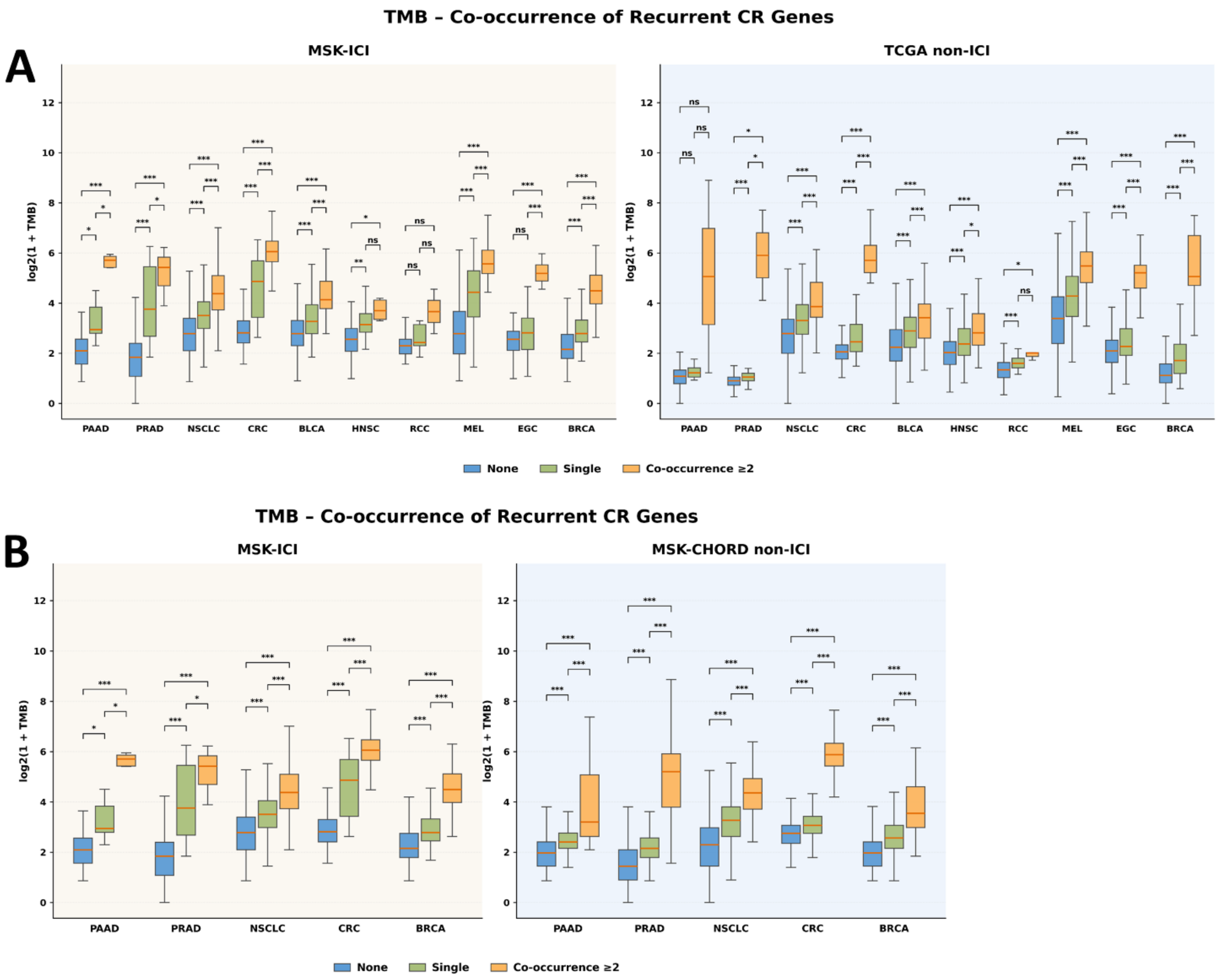


**Supplementary Figure 13. Tumor mutational burden according to co-occurrence of recurrent chromatin regulator mutations across ICI-treated and non-ICI-treated cohorts**. Boxplots show log2(1 + TMB) in tumors harboring no recurrent CR mutations, a single recurrent CR mutation, or co-occurring mutations in ≥2 of the six recurrent CR genes (*ARID1A, KMT2D, ARID1B, CREBBP, KMT2A,* and *EZH2*). (A) Comparisons between MSK-ICI and corresponding TCGA non-ICI cohorts across 10 cancer types. (B) Comparisons between MSK-ICI and MSK-CHORD non-ICI cohorts across PAAD, PRAD, NSCLC, CRC, and BRCA. Group differences were assessed using Kruskal–Wallis tests followed by Dunn's post-hoc comparisons. Blue, green, and orange denote no mutation, single mutation, and co-occurrence of ≥2 recurrent CR mutations, respectively. ns, not significant; *P < 0.05, **P < 0.01, ***P < 0.001.

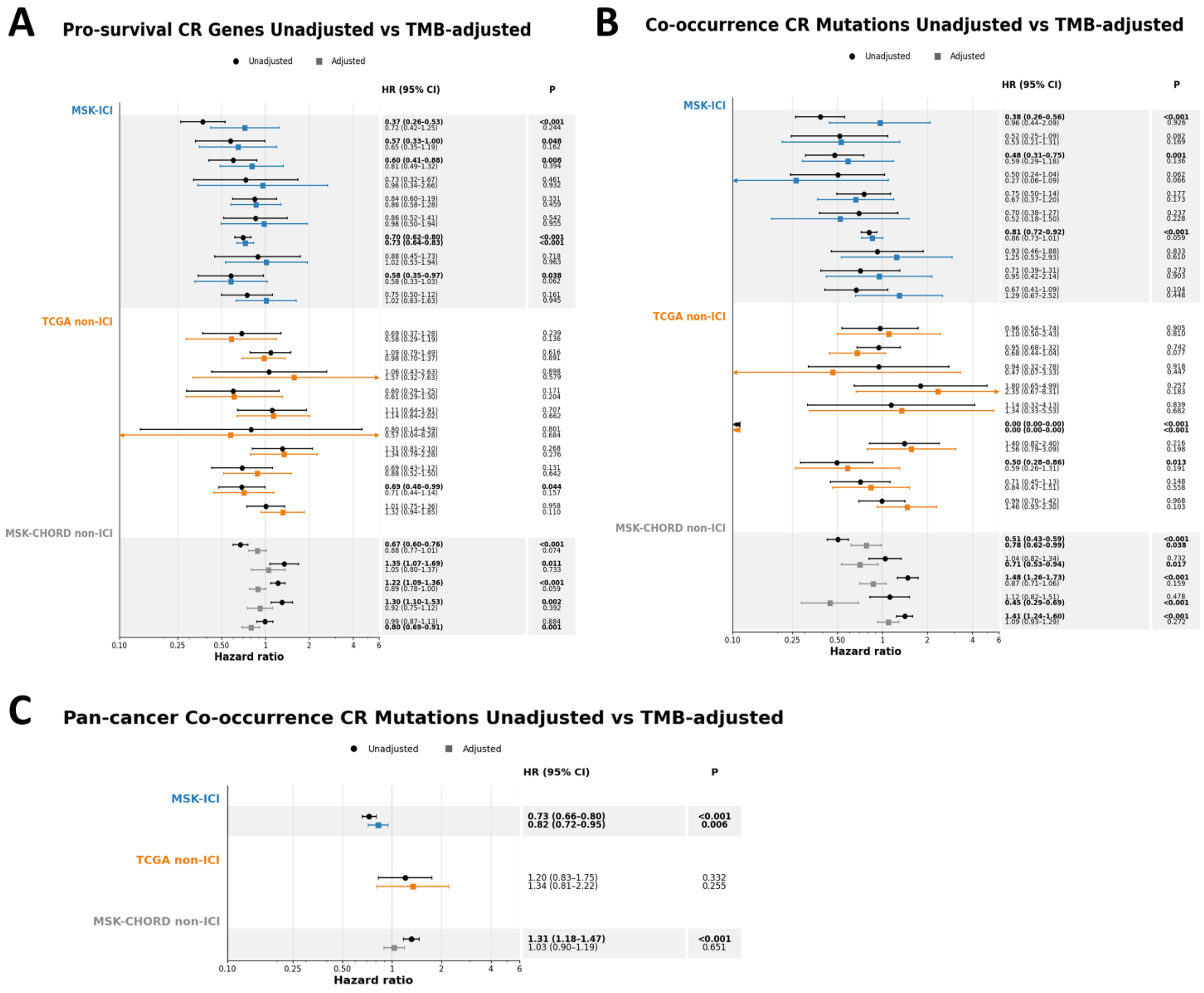


**Supplementary Data 14. Effect of tumor mutational burden adjustment on CR-associated survival estimates.** Forest plots compare unadjusted and TMB-adjusted ATT-IPTW-weighted Cox proportional-hazards models. (A) Cancer-specific survival associations for the pro-survival CR gene sets. (B) Cancer-specific survival associations of recurrent CR co-occurrence, comparing tumors harboring ≥2 recurrent CR mutations with tumors carrying a single recurrent CR mutation. (C) Corresponding pan-cancer analysis of recurrent CR co-occurrence in MSK-ICI, TCGA non-ICI, and MSK-CHORD non-ICI cohorts. Circles indicate unadjusted estimates and squares TMB-adjusted estimates; horizontal lines represent 95% confidence intervals (CIs). Exact hazard ratios (HRs), 95% CIs, and P values are shown on the right. HR < 1 indicates lower mortality hazard relative to the corresponding reference group.

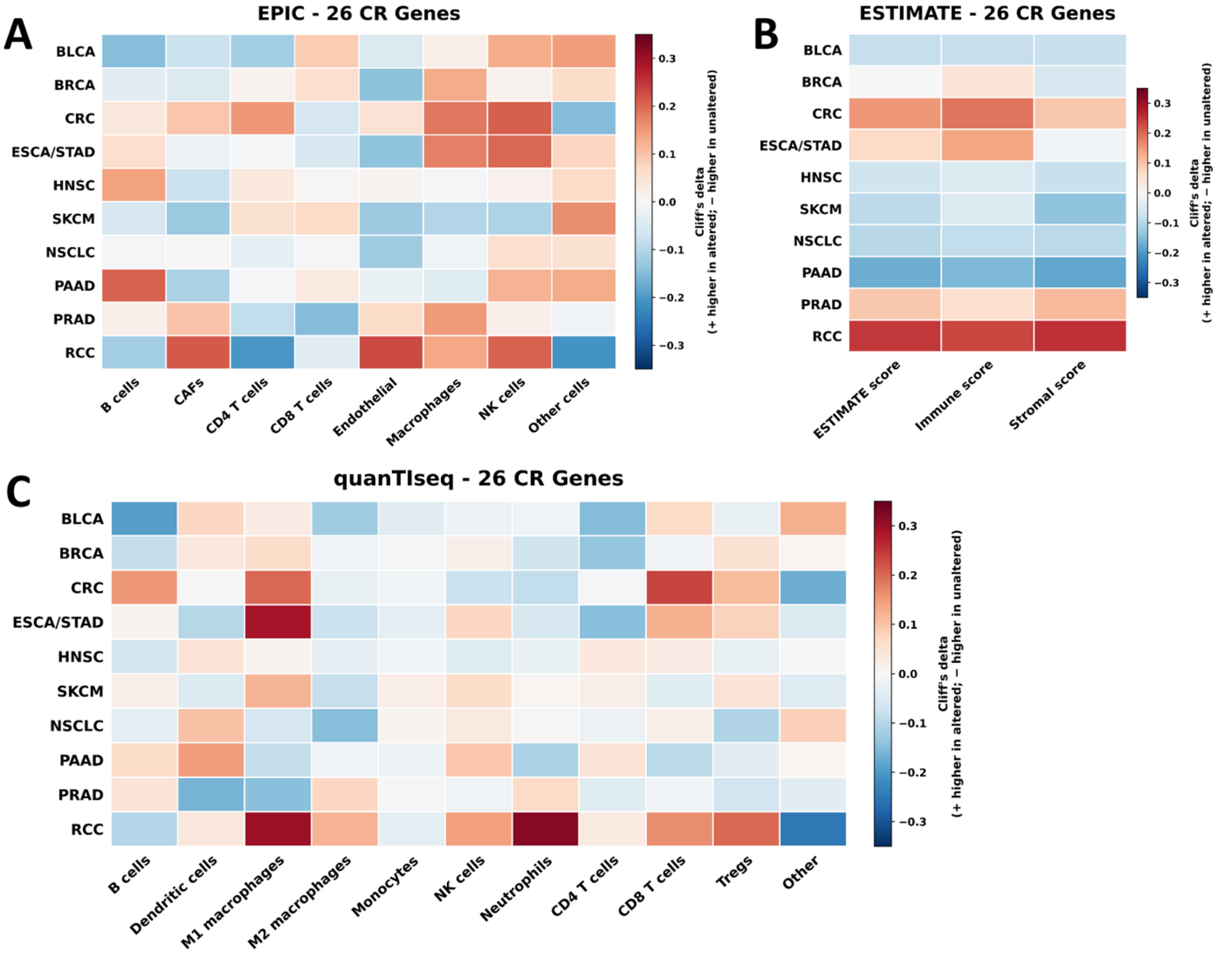


**Supplementary Figure 15. Tumor microenvironment differences between tumors altered and unaltered in the complete 26-CR gene set across cancer types.** Tumor microenvironment composition was estimated from TCGA bulk RNA-seq data by comparing tumors harboring mutations in at least one of the 26 chromatin regulator (CR) genes with tumors lacking these alterations. (A) EPIC-derived immune and stromal cell fractions. (B) ESTIMATE-derived immune, stromal, and combined ESTIMATE scores. (C) quanTIseq-derived immune-cell fractions. Heatmaps show Cliff's delta as the effect-size measure, with positive values indicating higher estimated cell fractions or scores in CR-altered tumors and negative values indicating higher values in CR-unaltered tumors. Rows represent cancer types and columns the indicated cell populations or ESTIMATE scores.

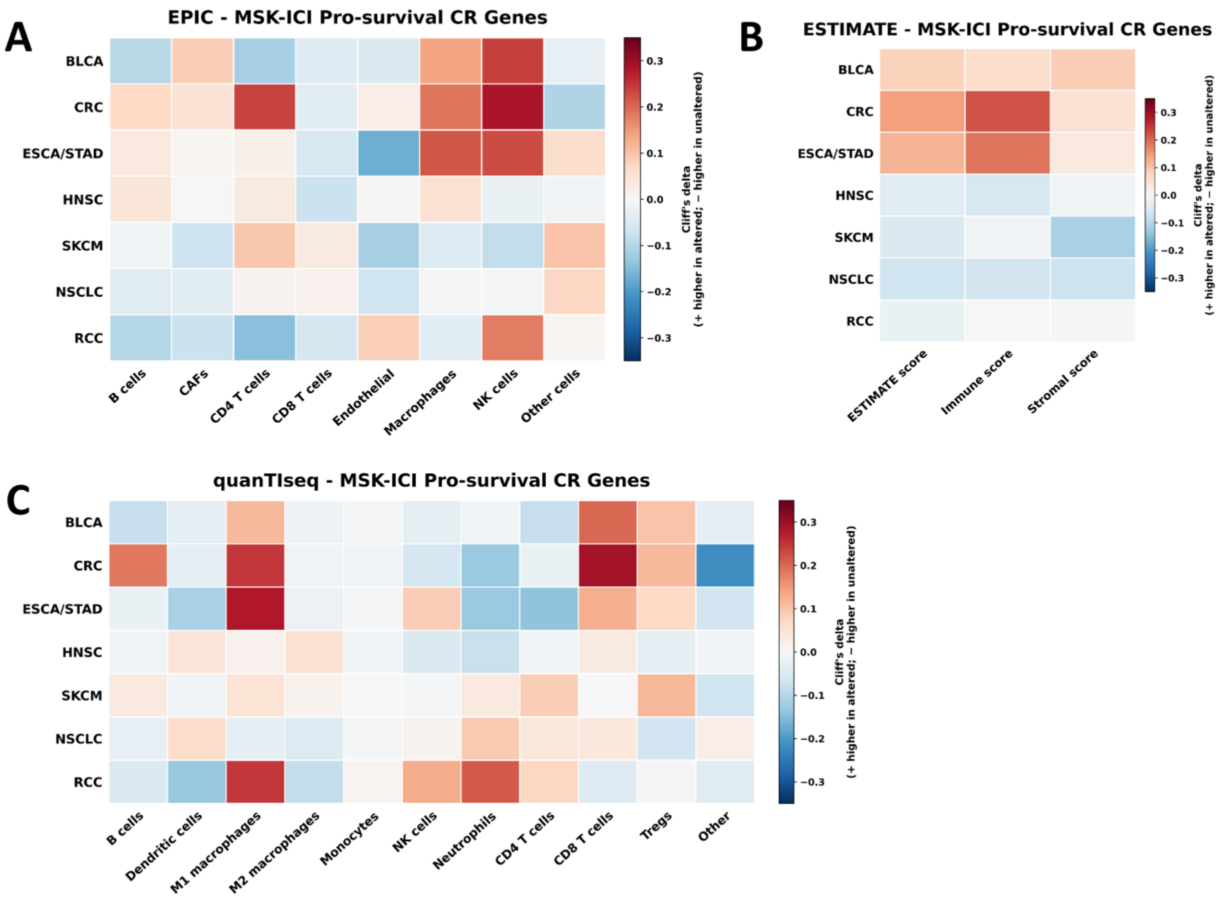


**Supplementary Figure 16. Tumor microenvironment differences between tumors altered and unaltered in any of the cancer-specific pro-survival genes across cancer types.** Tumor microenvironment composition was estimated from TCGA bulk RNA-seq data by comparing tumors harboring mutations in at least one of the cancer-specific pro-survival chromatin regulator (CR) genes with tumors lacking these alterations. (A) EPIC-derived immune and stromal cell fractions. (B) ESTIMATE-derived immune, stromal, and combined ESTIMATE scores. (C) quanTIseq-derived immune-cell fractions.  Heatmaps show Cliff's delta as the effect-size measure, with positive values indicating higher estimated cell fractions or scores in CR-altered tumors and negative values indicating higher values in CR-unaltered tumors. Rows represent cancer types and columns the indicated cell populations or ESTIMATE scores.

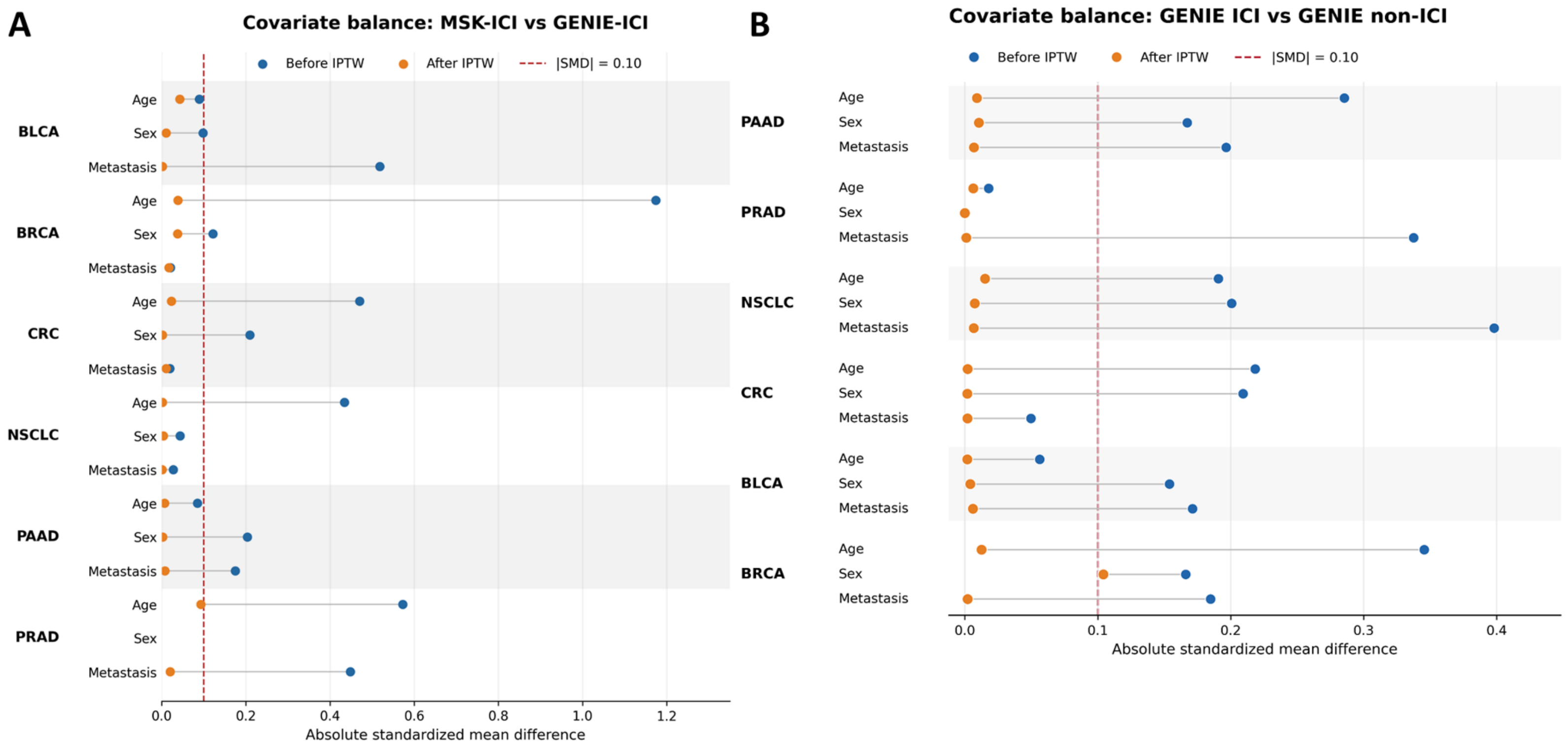


**Supplementary Figure 17. Covariate balance before and after inverse probability of treatment weighting in the GENIE validation analyses**. Love plots show absolute standardized mean differences (|SMD|) for age, sex, and metastatic status before and after ATT-IPTW across the indicated cancer types. (A) Balance between the MSK-ICI discovery cohort and GENIE-ICI cohort. (B) Balance between GENIE ICI and GENIE non-ICI cohorts. Blue points indicate pre-weighting and orange points post-weighting values; the dashed vertical line denotes the predefined threshold for adequate balance (|SMD| = 0.10). Missing covariate information is indicated where applicable.

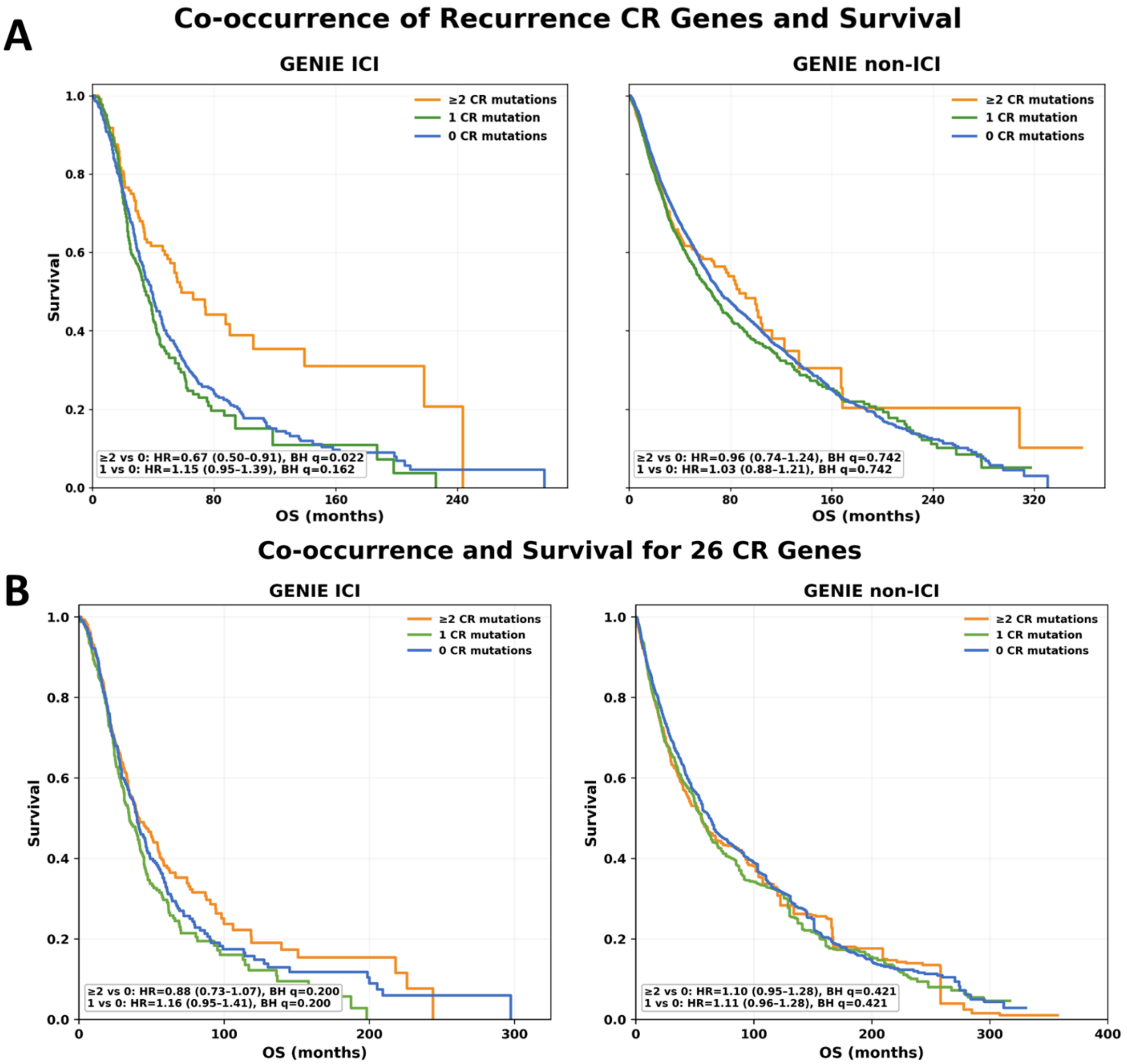


**Supplementary Figure 18. Survival associations of co-occurring chromatin regulator mutations in the AACR Project GENIE validation cohort**. IPTW-weighted Kaplan–Meier curves compare overall survival among tumors harboring ≥2, 1, or 0 CR mutations in GENIE ICI and GENIE non-ICI cohorts. (A) Co-occurrence within the six recurrent CR genes (*ARID1A, KMT2D, ARID1B, CREBBP, KMT2A,* and *EZH2*). (B) Co-occurrence across the complete 26-CR gene set. Pairwise hazard ratios (HRs), 95% confidence intervals (CIs), and BH-FDR-adjusted q-values are shown within each panel for ≥2 versus 0 mutations and 1 versus 0 mutations. Orange, green, and blue curves denote ≥2, 1, and 0 CR mutations, respectively.

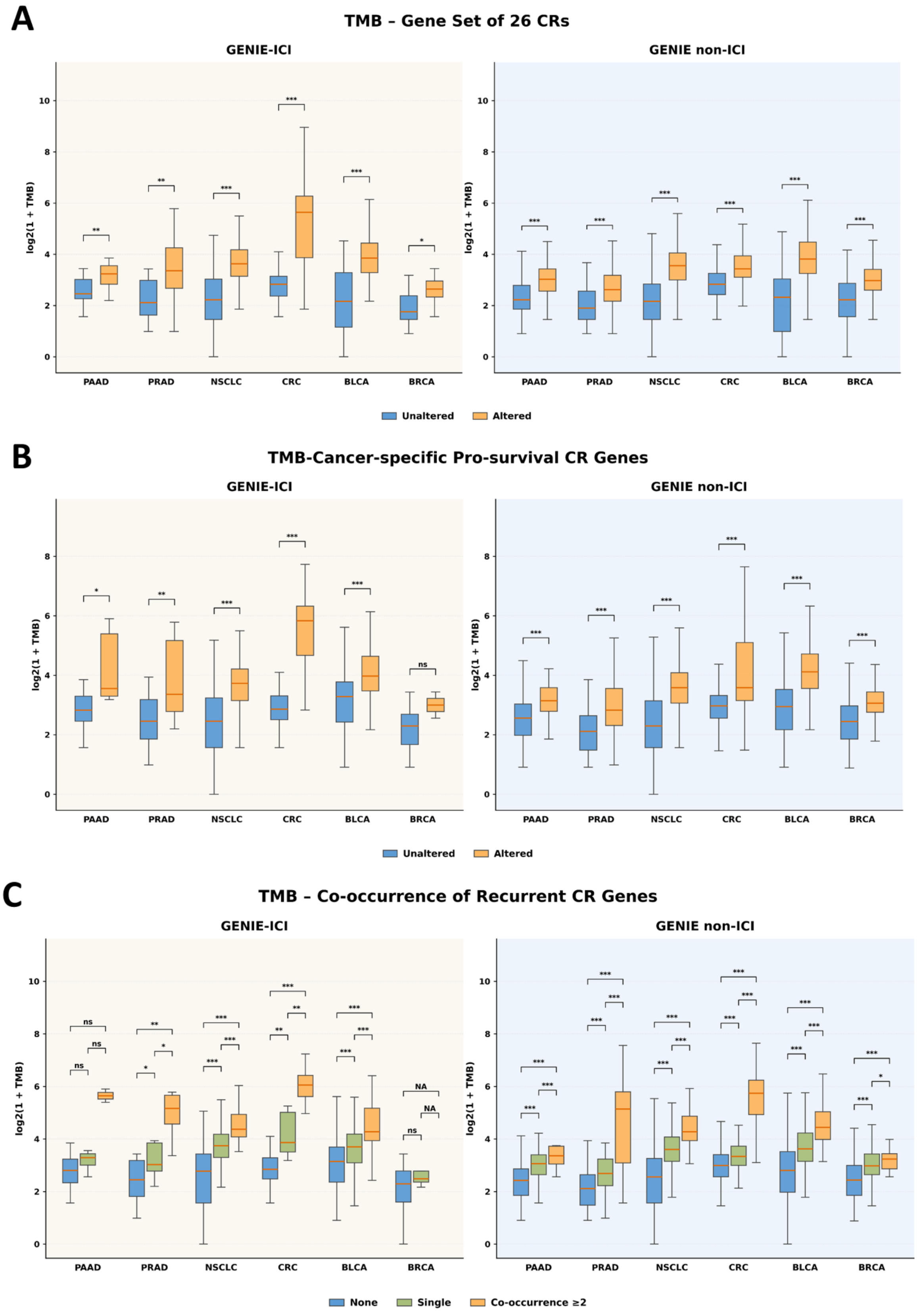
A
TMB – Gene Set of 26 CRs
GENIE-ICI
GENIE non-ICI
log2(1 + TMB)
PAAD
PRAD
NSCLC
CRC
BLCA
BRCA
Unaltered
Altered
B
TMB-Cancer-specific Pro-survival CR Genes
GENIE-ICI
GENIE non-ICI
log2(1 + TMB)
PAAD
PRAD
NSCLC
CRC
BLCA
BRCA
Unaltered
Altered
C
TMB – Co-occurrence of Recurrent CR Genes
GENIE-ICI
GENIE non-ICI
log2(1 + TMB)
PAAD
PRAD
NSCLC
CRC
BLCA
BRCA
None
Single
Co-occurrence ≥2

**Supplementary Figure 19. Validation of the association between chromatin regulator alterations and tumor mutational burden in AACR Project GENIE. Boxplots show log2(1 + TMB) across six cancer types in GENIE ICI and GENIE non-ICI cohorts**. (A) TMB in tumors altered versus unaltered in the complete 26-CR gene set. (B) TMB in tumors altered versus unaltered in the cancer-specific pro-survival CR gene sets defined in the MSK-ICI discovery cohort. (C) TMB according to recurrent CR mutation burden, comparing tumors with no recurrent CR mutations, a single mutation, or co-occurring mutations in ≥2 recurrent CR genes. Cancer types include pancreatic adenocarcinoma (PAAD), prostate adenocarcinoma (PRAD), non-small cell lung cancer (NSCLC), colorectal cancer (CRC), bladder cancer (BLCA), and breast cancer (BRCA). Two-group comparisons were assessed using two-sided Mann–Whitney U tests, whereas three-group comparisons used Kruskal–Wallis tests followed by Dunn's post-hoc comparisons. ns, not significant; *P < 0.05, **P < 0.01, ***P < 0.001; NA, comparison not available.
